# Natural killer cell educating KIR/HLA combinations impact survival in anti-PD-L1 treated cancer patients

**DOI:** 10.1101/2022.12.06.22282592

**Authors:** David Roe, Howard Rosoff, Dan Fu Ruan, Zia Khan, Pranay Dogra, Jonathan Carroll, Julie Hunkapiller, Rajat Mohindra, Minu K. Srivastava, Barzin Y. Nabet, G. Scott Chandler, Matthew L. Albert, Mark I. McCarthy, Ira Mellman, Amir Horowitz, Christian Hammer

## Abstract

Natural killer (NK) cells are educated through the binding of killer immunoglobulin like receptors (KIR) to human leukocyte antigen (HLA) proteins, but it is unknown whether the presence of these highly diverse KIR/HLA interactions influence responses to immunotherapy in solid tumors. We report herein two observations that shed light on NK cell function and abundance in anti-tumor immune responses. In patients with non-small cell lung cancer treated with anti-PD-L1 therapy, we found that individuals carrying HLA-C1 and HLA-Bw4 alleles and the genes coding for their receptors KIR2DL3 and KIR3DL1 showed improved overall survival (OS). Combined with our second finding that NK cell infiltration was independently associated with improved OS, our findings have important implications for precision medicine approaches and the development of NK cell-based therapies.

## Main text

Cancer immunotherapy has revolutionized cancer treatment, but the large variability in patient responses calls for novel treatment strategies and biomarker-based enrichment of responders. Human leukocyte antigen (HLA) class I molecules present antigenic peptides on tumor cells to CD8^+^ T cells as a key component of what is known as signal 1 in immunology, but they also have core functions in the signaling processes of other immune cell types (1). They play a key role in the education of natural killer (NK) cells, which have been proposed as important players in innate anti-tumor immune responses. NK cells have direct cytotoxic activity, being able to induce both apoptosis and pyroptosis in target cells, but they can also influence the inflammatory environment via the secretion of cytokines and chemokines (2). NK cell functions are well investigated in mouse models, and correlative studies have also defined an important role in human cancers (2, 3). Several studies showed improved survival in patients with higher NK cell infiltration in the tumor (2, 4, 5), and NK cells also express immune checkpoints such as PD-1, such that PD-L1 expression on tumor cells can contribute to their inhibition. However, these studies are correlative in nature and cannot establish causal relationships.

NK cell education is the important process by which the cells acquire effector function. They are educated via two molecular mechanisms, mediated by the interaction of NKG2A and HLA-E, and their respective engagement of highly polymorphic HLA class I molecules with Killer Immunoglobulin like receptors (KIR) (6, 7). HLA class I alleles can be classified into ligand groups based on amino acid residues that determine their interactions with specific inhibitory KIR (Figure 1A). Importantly, these KIR are expressed on CD56^dim^ NK cells, which are relatively abundant in blood, spleen and lung tissue of healthy individuals, and display significantly higher cytotoxic activity than CD56^bright^ NK cells, which do not typically express KIR (8, 9). Different models for NK cell education have been proposed, but all of them are compatible with the idea of ‘tuning’ of NK cell reactivity as a response to environmental changes. These models are based on the interaction of HLA class I molecules with inhibitory receptors on NK cells, the presence of such interactions being associated with higher effector function (10). This is of relevance to the concept of ‘missing-self’ recognition. Tumors commonly display reduced or abolished HLA class I expression, and NK cells can detect this as a stress signal and target the stressed cell (1). There are also potential implications for the role of NK cell education in the context of anti-PD-1 / anti-PD-L1 checkpoint blockade. PD-1 is usually not highly expressed on NK cells of healthy individuals, but has been found on peripheral and intratumoral NK cells of cancer patients, mainly associated with the CD56^dim^ phenotype (11–13). In an investigation in non-small cell lung cancer (NSCLC) patients, a subset of tumor-infiltrating NK cells expressed PD-1. These cells were found to be more dysfunctional and to express more inhibitory receptors, but PD-1 blockade was able to restore their functionality (14).

**Fig. 1.**
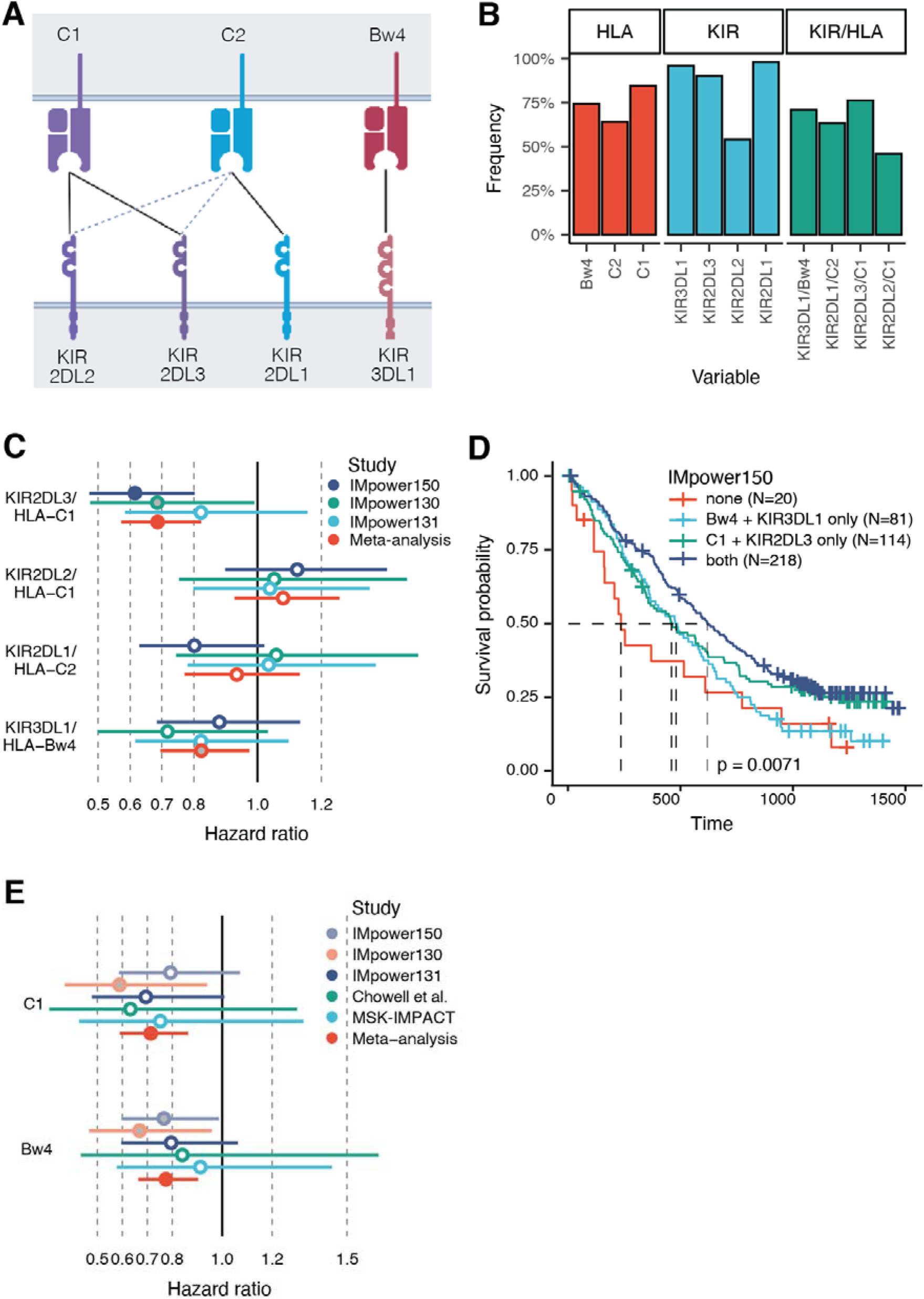
Impact of KIR/HLA combinations and HLA ligand status on overall survival in NSCLC patients. (**A**) High-frequency KIR/HLA combinations according to Pende et al. (41). HLA class I alleles are grouped into C1, C2 and Bw4 ligand groups and interact with specific KIR on NK cells. Dashed lines depict low-affinity interactions, which are not considered in this study. (**B**) Prevalence of HLA ligand groups, KIR genes, and KIR/HLA combinations in IMpower150 patients. Reference and other trial frequencies are provided in Table S1. (**C**) Association between KIR/HLA combinations and overall survival for patients with NSCLC treated with atezolizumab in IMpower150 (N=433), IMpower130 (N=202) and IMpower131 (N=319), as well as meta-analysis results (N=954). Color-filled circles represent statistical significance after adjustment for multiple testing, and gray-filled circles nominal significance. Horizontal lines depict 95% confidence intervals. The complete data and results for control arms of the trials are provided in Table S3. (**D**) Kaplan-Meier estimates of overall survival in IMpower150 for atezolizumab-treated patients carrying none of the two significant KIR/HLA interactions KIR3DL1-Bw4 and KIR2DL3/HLA-C1 (N=20), one of the two (N=81 and N=114, respectively), and both (N=218). Dashed lines depict median survival for each group. P-value is based on a log-rank test. (**E**) Association between HLA ligand groups C1 and Bw4, and overall survival for patients with NSCLC treated with immune checkpoint inhibitors. Included cohorts are the atezolizumab clinical trials IMpower150 (N=433), IMpower130 (N=202) and IMpower131 (N=319), and two replication datasets (Chowell et al., N= 101; MSK-IMPACT, N=271). Meta-analysis results are based on a total of 1326 patients. Color-filled circles represent statistical significance after adjustment for multiple testing, and gray-filled circles nominal significance. Horizontal lines depict 95% confidence intervals. The complete data and results for control arms of the trials are provided in Table S5.

We hypothesized that survival in patients with NSCLC treated with immune checkpoint inhibitors might be impacted by the presence or absence of NK-cell educating KIR-HLA combinations. We performed germline whole-genome sequencing (WGS) in a discovery cohort (IMpower150) of 433 patients of European ancestry with metastatic non-squamous NSCLC treated with the PD-L1 antibody atezolizumab in combination with chemotherapy, and 182 patients in the control arms of this study (Suppl. Methods, Table S1) (15). Applying validated methods, HLA alleles and KIR gene status were computationally inferred, and variables defining the presence of four common KIR-HLA combinations that were shown to have a significant contribution to NK cell education were computed (Suppl. Methods) (16). All observed frequencies were in the expected range when compared to published reference data for European populations (Figure 1B, Table S2).

Median overall survival was significantly improved in patients treated with atezolizumab who carried HLA-C1 alleles and their receptor *KIR2DL3*, when compared to patients who did not carry both ligand and receptor [5.1 months, P < 3.4×10^−4^, hazard ratio (HR) = 0.62]. No other variables showed a significant association after adjustment for multiple testing, and none reached statistical significance in the control arm of the study (Table S3).

To replicate this finding and to improve statistical power, we inferred HLA and KIR types in two other NSCLC trials using combinations of atezolizumab and chemotherapy, IMpower130 (non-squamous NSCLC) and IMpower131 (squamous NSCLC) (Table S1) (17, 18). In a meta-analysis, the *KIR2DL3/*HLA-C1 association was confirmed in a total of 954 patients of European ancestry who received atezolizumab [P_meta_ = 5.0×10^−5^, HR=0.69], with highly consistent effect estimates across the three trials (Figure 1C, Table S4). In addition, the interaction of *KIR3DL1* and HLA-Bw4 was nominally associated with improved OS [P_meta_= 0.02, HR=0.82] (Figure 1C, Table S4). Again, no significant associations were found for the control arms of the studies (total N=440, Figure S1, Table S4), which is why we focused on atezolizumab treated patients only from this point.

Previous disease association studies of allelic variation in *KIR2D* genes and *HLA-C* may shed some light on the question of why these combinations showed significant associations. Carriers of the *KIR A* haplotype encode strongly inhibitory KIR2DL1 allotypes relative to KIR2DL1 encoded on *KIR B* haplotypes (19). Conversely, *KIR A*-encoded KIR2DL3 allotypes bind with weaker affinity and lower inhibitory strength compared to the *KIR-B*-encoded KIR2DL2. These features frame a ‘rheostat’ model of potential hierarchy in inhibitory strength across the family of KIR receptors (KIR2DL1/HLA-C2 > KIR2DL2/HLA-C1 > KIR2DL3/HLA-C1) that when expressed in combination on NK cells results in ‘tuning’ of sensitivity to changes in HLA class I expression on neighboring cells (20). This model when applied to NSCLC positions KIR2DL3 as the weaker inhibitory checkpoint for NK cells and might suggest a lower threshold for concomitant signaling through activating NK cell receptors. Relative strength in binding by and inhibition through KIR3DL1 interactions with HLA-Bw4 molecules have been studied as well, where different combinations vary significantly in cell surface expression and in their strength of interactions (21–23). The effects of these interactions stratify response to microbial infections (24), risk of susceptibility to autoimmunity (25), and relapse of acute myeloid leukemia (AML) following hematopoietic cell transplantation (26).

Since NK cells are educated by multiple KIR/HLA interactions, we hypothesized that the presence of both *KIR2DL3/*HLA-C1 and *KIR3DL1/*HLA-Bw4 might result in improved outcome when compared to groups of patients with only one or none of the two combinations. In the IMpower150 discovery cohort, we indeed observed the strongest separation in a comparison of groups of patients with both vs. none (Figure 1D), with a median OS difference of 13.2 months. This was consistent in both IMpower130 and IMpower131 (Fig. S2, pairwise comparisons in Table S5), resulting in a strong association in a meta-analysis comparing these two groups [P_meta_ = 0.002, HR = 0.59]. Our finding is also consistent with the rheostat model of NK cell education, which proposes that the effector potential of NK cells positively correlates with the number of HLA class I interactions with inhibitory receptors (10).

Based on these findings, we also evaluated whether HLA ligand groups alone, independent of the presence of their respective KIR, showed significant associations in meta-analyses. In addition to our three atezolizumab clinical trials, we included NSCLC patients from two cancer immunotherapy cohorts with available HLA genotypes previously investigated by Chowell et al. (27), analyzing a total of 1326 patients who received immune checkpoint blockade (ICB). Both HLA-C1 [P_meta_ = 4.9×10^−4^, HR = 0.71] and HLA-Bw4 [P_meta_ = 1.1×10^−3^, HR = 0.77] carrier status were associated with longer OS (Figure 1E, Table S6). This is likely due to the fact that less than 10% of genotyped patients did not carry *KIR2DL3*, and less than 5% did not carry *KIR3DL1* (Table S2). Most patients with HLA-C1 or HLA-Bw4 alleles therefore have NK cells educated via the associated interactions. KIR gene carrier status alone was not significantly associated with OS across these studies, although *KIR2DL3* carriers showed significantly longer OS in the IMpower150 discovery cohort only (Table S7).

Single HLA alleles or supertypes were previously reported to be associated with outcomes in patients treated with ICB (27, 28), but the results were not consistent across studies and could not be replicated in large follow-up analyses (29, 30). We therefore wanted to exclude that our results might be driven by strong associations of single HLA variants that belong to the HLA-C1 or HLA-Bw4 ligand groups. Indeed, no single allele or supertype was found to be associated with OS, and none of the previously associated variables achieved nominal significance (Tables S8 and S9). While a patient’s HLA genotype defines the spectrum of antigens that can be presented on tumor cells, every tumor has a unique mutational profile, which is in contrast to, for example, autoimmune diseases, where HLA allelic associations are abundant and at least in some cases related to disease-specific self-antigens. This increased complexity might explain the lack of allele-specific associations.

It was also previously hypothesized that HLA class I heterozygosity or evolutionary divergence might be associated with improved outcomes. Both should overall result in a larger spectrum of antigens that can be presented by an individual’s pool of HLA alleles, which might in turn increase the likelihood for a potent anti-tumor immune response. Chowell et al. showed significant associations with both heterozygosity and divergence (Grantham distance) (27, 31), but their results could not be replicated in more recent analyses (29, 30). In our analysis, no results were significant after adjustment for multiple testing, however *HLA-C* evolutionary divergence was nominally significantly associated with longer OS [p=0.04, Beta=0.98] (Tables S10 and S11). While the hypothesis is conceptually sound, the breadth of anti-tumor immune responses is not yet understood in sufficient detail, and phenomena such as immunodominance might limit the theoretical benefits of a larger pool of presented neoantigens (32).

In addition to NK cell reactivity, their infiltration into tumor tissue could be an important determinant of patient outcomes. Cursons et al. previously used published NK cell signatures to curate a gene set that estimates the abundance of NK cells in a tumor(4). They demonstrated that a gene score derived from this set was associated with improved survival in melanoma patients, using RNA sequencing (RNA-seq) data from The Cancer Genome Atlas (TCGA) (4, 33). We applied the same gene signature to the IMpower150 study, using RNA-seq data from tumor biopsies. First, we examined whether the relevant KIR-HLA combinations were associated with this NK cell score, in which case the genetic associations might be explained by increased NK cell infiltration, still possibly due to better education. However, we did not find significant differences between patients with and without the *KIR2DL3*/HLA-C1 and *KIR3DL1*/HLA-Bw4 combinations, respectively (Figure 2A).

**Fig. 2.**
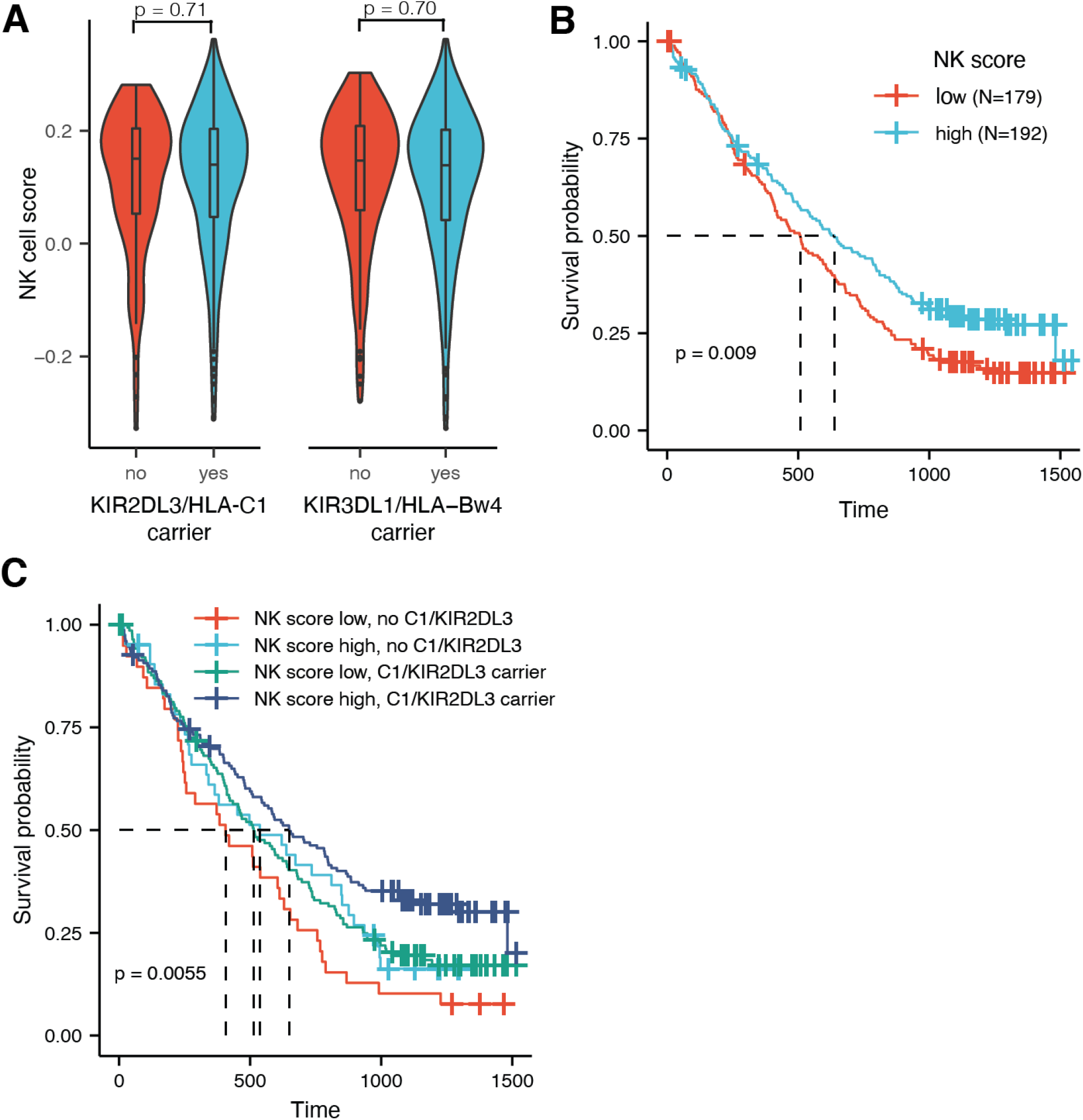
Impact of NK cell infiltration alone and in combination with KIR/HLA status on overall survival in IMpower150. (**A**) Violin plots representing the distribution of the NK cell infiltration signature in carriers vs. non-carriers of the two associated KIR-HLA combinations. Box plots depict median and interquartile range. No significant differences were found. (**B**) Kaplan-Meier estimates of overall survival for atezolizumab-treated patients with low (below median, N=179) and high (above median, N=192) NK cell score. Dashed lines depict median survival for each group. P-value is based on a log-rank test. (**C**) Kaplan-Meier estimates of overall survival for atezolizumab-treated patients according to NK cell score and presence of the KIR2DL3/HLA-C1 combination. Dashed lines depict median survival for each group. P-value is based on a log-rank test.

In 371 patients treated with atezolizumab, an above median NK cell score was associated with longer OS [4.3 months, P = 0.008, HR = 0.73] (Figure 2B). No significant association was found for the control arms [N = 151, P = 0.49, HR = 0.88] (Figure S3). While these results suggest a role of NK cell abundance in outcome following ICB (‘quantity’), the associations of KIR/HLA combinations point to the importance of NK cell education (‘quality’). We therefore evaluated whether infiltration and education independently impact OS in patients treated with atezolizumab, using a multivariable model including the NK cell score and the four previously tested KIR/HLA combinations. Both the NK cell score [P = 0.015, HR = 0.74] and the presence of *KIR2DL3/*HLA-C1 [P = 0.012, HR = 0.69] were significantly associated with longer OS in this model (Table S12), but none of the other KIR/HLA combinations, which is consistent with the previous analysis of KIR/HLA combinations in IMpower150 (Table S3). Patients who had a high NK cell score and carried *KIR2DL3/*HLA-C1 showed the best overall survival, significantly different and 8 months longer compared to patients with low NK cell infiltration and without the KIR/HLA combination (Figure 2C, Table S13). Patients with only high NK cell score or the KIR/HLA combination showed intermediate OS (Figure 2C), but there were no statistically significant differences after adjustment for multiple testing, possibly due to limited statistical power (Table S13).

In summary, our results demonstrate an important role of NK cells in anti-tumor immune responses in patients treated with cancer immunotherapy. In addition to their abundance in the tumor microenvironment (TME), we showed that their education via germline-encoded KIR/HLA interactions impacts overall survival in atezolizumab-treated NSCLC patients. These findings, based on human genetics and thus establishing a causal relationship, might have implications for clinical trial design, and in combination with other tumor intrinsic and extrinsic factors contribute to statistical models better predicting patient outcomes. Development of NK cell therapies for cancer will also benefit from consideration of how NK cells are educated and whether HLA-A,-B,-C and HLA-E expression are regulated similarly or differently by tumors in the TME. Finally, larger follow-up analyses will be required to conclusively answer questions related to the importance of single HLA alleles and HLA allelic divergence.

## Data Availability

Qualified researchers may request access to individual patient data used in this study through Roche's data sharing platforms in accordance with the Global Policy on Sharing of Clinical Study Information:
http://www.roche.com/research_and_development/who_we_are_how_we_work/clinical_trials/our_commitment_to_data_sharing.htm. To ensure compliance with legal, data retention, and patient confidentiality obligations in the informed consent forms (ICF), the whole-genome sequencing data collected (in VCF or BAM/FASTQ formats) cannot be hosted on a public, controlled access repository and will be made available to individual requesters on completion of a data sharing agreement with Roche/Genentech. Requests for access to whole-genome sequencing data should be made to the corresponding author by email at. The planned research with the requested data will be reviewed by the Roche Pharma Repository Governance Committee to assess its scientific merit and to ensure it is within scope of the ICF approved locally at each study site.

## Acknowledgements

We thank all of our Genentech colleagues involved in the Human Genetics Initiative including Natalie Bowers, Jens Reeder, and Suresh Selvaraj. We acknowledge the Roche-wide Enhanced Data and Insights Sharing (EDIS) network. We also thank the IMpower150, IMpower130, and IMpower131 study teams, the investigators, and the patients that contributed to this study.

## Author contributions

Conceptualization: AH, CH

Methodology: DR, ZK, JC, JH, RM, MKS, BYN, GSC

Investigation: DR, HR, DFR, PD, CH

Visualization: DR, CH

Supervision: MLA, MM, IM, CH

Writing - original draft: DR, CH

Writing - review & editing: DR, HR, DFR, ZK, PD, JC, JH, RM, MKS, BYN, GSC, MIM, IM, AH, CH

## Data and materials availability

Qualified researchers may request access to individual patient data used in this study through Roche’s data sharing platforms in accordance with the Global Policy on Sharing of Clinical Study Information: http://www.roche.com/research_and_development/who_we_are_how_we_work/clinical_trials/our_commitment_to_data_sharing.htm.

To ensure compliance with legal, data retention, and patient confidentiality obligations in the informed consent forms (ICF), the whole-genome sequencing data collected (in VCF or BAM/FASTQ formats) cannot be hosted on a public, controlled access repository and will be made available to individual requesters on completion of a data sharing agreement with Roche/Genentech. Requests for access to whole-genome sequencing data should be made to the corresponding author by email at. The planned research with the requested data will be reviewed by the Roche Pharma Repository Governance Committee to assess its scientific merit and to ensure it is within scope of the ICF approved locally at each study site.

## Supplementary Materials

Materials and Methods

Figs. S1 - S3

Tables S1 - S12

Data S1

## References

1. S. Jhunjhunwala, C. Hammer, L. Delamarre, Antigen presentation in cancer: insights into tumour immunogenicity and immune evasion. Nat Rev Cancer, 1–15 (2021).

2. N. K. Wolf, D. U. Kissiov, D. H. Raulet, Roles of natural killer cells in immunity to cancer, and applications to immunotherapy. Nat Rev Immunol, 1–16 (2022).

3. N. Shimasaki, A. Jain, D. Campana, NK cells for cancer immunotherapy. Nat Rev Drug Discov. 19, 200–218 (2020).

4. J. Cursons, F. Souza-Fonseca-Guimaraes, M. Foroutan, A. Anderson, F. Hollande, S. Hediyeh-Zadeh, A. Behren, N. D. Huntington, M. J. Davis, A Gene Signature Predicting Natural Killer Cell Infiltration and Improved Survival in Melanoma Patients. Cancer Immunol Res. 7, 1162–1174 (2019).

5. S. Nersesian, S. L. Schwartz, S. R. Grantham, L. K. MacLean, S. N. Lee, M. Pugh-Toole, J. E. Boudreau, NK cell infiltration is associated with improved overall survival in solid cancers: A systematic review and meta-analysis. Transl Oncol. 14, 100930 (2021).

6. A. Horowitz, Z. Djaoud, N. Nemat-Gorgani, J. Blokhuis, H. G. Hilton, V. Béziat, K.-J. Malmberg, P. J. Norman, L. A. Guethlein, P. Parham, Class I HLA haplotypes form two schools that educate NK cells in different ways. Sci Immunol. 1, eaag1672–eaag1672 (2016).

7. P. Parham, L. A. Guethlein, Genetics of Natural Killer Cells in Human Health, Disease, and Survival. Annu Rev Immunol. 36, 1–30 (2018).

8. P. Dogra, C. Rancan, W. Ma, M. Toth, T. Senda, D. J. Carpenter, M. Kubota, R. Matsumoto, P. Thapa, P. A. Szabo, M. M. L. Poon, J. Li, J. Arakawa-Hoyt, Y. Shen, L. Fong, L. L. Lanier, D. L. Farber, Tissue Determinants of Human NK Cell Development, Function, and Residence. Cell. 180, 749-763.e13 (2020).

9. N. K. Björkström, H.-G. Ljunggren, J. Michaëlsson, Emerging insights into natural killer cells in human peripheral tissues. Nat Rev Immunol. 16, 310–320 (2016).

10. J. E. Boudreau, K. C. Hsu, Natural Killer Cell Education and the Response to Infection and Cancer Therapy: Stay Tuned. Trends Immunol. 39, 222–239 (2018).

11. L. Quatrini, F. R. Mariotti, E. Munari, N. Tumino, P. Vacca, L. Moretta, The Immune Checkpoint PD-1 in Natural Killer Cells: Expression, Function and Targeting in Tumour Immunotherapy. Cancers. 12, 3285 (2020).

12. A. K. Wagner, N. Kadri, C. Tibbitt, K. van de Ven, S. Bagawath-Singh, D. Oliinyk, E. LeGresly, N. Campbell, S. Trittel, P. Riese, T. Sandalova, A. Achour, K. Kärre, B. J. Chambers, Biorxiv, in press, doi:10.1101/2021.03.29.437486.

13. Z. Davis, M. Felices, T. Lenvik, S. Badal, J. T. Walker, P. Hinderlie, J. L. Riley, D. A. Vallera, B. R. Blazar, J. S. Miller, Low-density PD-1 expression on resting human natural killer cells is functional and upregulated after transplantation. Blood Adv. 5, 1069–1080 (2021).

14. M. P. Trefny, M. Kaiser, M. A. Stanczak, P. Herzig, S. Savic, M. Wiese, D. Lardinois, H. Läubli, F. Uhlenbrock, A. Zippelius, PD-1+ natural killer cells in human non-small cell lung cancer can be activated by PD-1/PD-L1 blockade. Cancer Immunol Immunother Cii, 1–13 (2020).

15. M. A. Socinski, R. M. Jotte, F. Cappuzzo, F. Orlandi, D. Stroyakovskiy, N. Nogami, D. Rodríguez-Abreu, D. Moro-Sibilot, C. A. Thomas, F. Barlesi, G. Finley, C. Kelsch, A. Lee, S. Coleman, Y. Deng, Y. Shen, M. Kowanetz, A. Lopez-Chavez, A. Sandler, M. Reck, Im. S. Group, Atezolizumab for First-Line Treatment of Metastatic Nonsquamous NSCLC. New Engl J Med. 378, 2288–2301 (2018).

16. J. E. Boudreau, K. C. Hsu, Natural killer cell education in human health and disease. Curr Opin Immunol. 50, 102–111 (2018).

17. H. West, M. McCleod, M. Hussein, A. Morabito, A. Rittmeyer, H. J. Conter, H.-G. Kopp, D. Daniel, S. McCune, T. Mekhail, A. Zer, N. Reinmuth, A. Sadiq, A. Sandler, W. Lin, T. O. Lohmann, V. Archer, L. Wang, M. Kowanetz, F. Cappuzzo, Atezolizumab in combination with carboplatin plus nab-paclitaxel chemotherapy compared with chemotherapy alone as first-line treatment for metastatic non-squamous non-small-cell lung cancer (IMpower130): a multicentre, randomised, open-label, phase 3 trial. Lancet Oncol. 20, 924–937 (2019).

18. R. Jotte, F. Cappuzzo, I. Vynnychenko, D. Stroyakovskiy, D. Rodríguez-Abreu, M. Hussein, R. Soo, H. J. Conter, T. Kozuki, K.-C. Huang, V. Graupner, S. W. Sun, T. Hoang, H. Jessop, M. McCleland, M. Ballinger, A. Sandler, M. A. Socinski, Atezolizumab in Combination With Carboplatin and Nab-Paclitaxel in Advanced Squamous Non-Small-Cell Lung Cancer (IMpower131): Results From a Randomized Phase III Trial. J Thorac Oncol. 15, 1351–1360 (2020).

19. H. G. Hilton, L. A. Guethlein, A. Goyos, N. Nemat-Gorgani, D. A. Bushnell, P. J. Norman, P. Parham, Polymorphic HLA-C Receptors Balance the Functional Characteristics of KIR Haplotypes. J Immunol. 195, 3160–3170 (2015).

20. P. Brodin, K. Kärre, P. Höglund, NK cell education: not an on-off switch but a tunable rheostat. Trends Immunol. 30, 143–149 (2009).

21. S. Kim, J. B. Sunwoo, L. Yang, T. Choi, Y.-J. Song, A. R. French, A. Vlahiotis, J. F. Piccirillo, M. Cella, M. Colonna, T. Mohanakumar, K. C. Hsu, B. Dupont, W. M. Yokoyama, HLA alleles determine differences in human natural killer cell responsiveness and potency. Proc National Acad Sci. 105, 3053–3058 (2008).

22. J. E. Gumperz, V. Litwin, J. H. Phillips, L. L. Lanier, P. Parham, The Bw4 public epitope of HLA-B molecules confers reactivity with natural killer cell clones that express NKB1, a putative HLA receptor. J Exp Medicine. 181, 1133–1144 (1995).

23. J. E. Boudreau, T. J. Mulrooney, J.-B. L. Luduec, E. Barker, K. C. Hsu, KIR3DL1 and HLA-B Density and Binding Calibrate NK Education and Response to HIV. J Immunol. 196, 3398– 3410 (2016).

24. S. Rajagopalan, E. O. Long, Understanding how combinations of HLA and KIR genes influence disease. J Exp Medicine. 201, 1025–1029 (2005).

25. G. W. Nelson, M. P. Martin, D. Gladman, J. Wade, J. Trowsdale, M. Carrington, Cutting Edge: Heterozygote Advantage in Autoimmune Disease: Hierarchy of Protection/Susceptibility Conferred by HLA and Killer Ig-Like Receptor Combinations in Psoriatic Arthritis. J Immunol. 173, 4273–4276 (2004).

26. S. Cooley, D. J. Weisdorf, L. A. Guethlein, J. P. Klein, T. Wang, C. T. Le, S. G. E. Marsh, D. Geraghty, S. Spellman, M. D. Haagenson, M. Ladner, E. Trachtenberg, P. Parham, J. S. Miller, Donor selection for natural killer cell receptor genes leads to superior survival after unrelated transplantation for acute myelogenous leukemia. Blood. 116, 2411–2419 (2010).

27. D. Chowell, L. G. T. Morris, C. M. Grigg, J. K. Weber, R. M. Samstein, V. Makarov, F. Kuo, S. M. Kendall, D. Requena, N. Riaz, B. Greenbaum, J. Carroll, E. Garon, D. M. Hyman, A. Zehir, D. Solit, M. Berger, R. Zhou, N. A. Rizvi, T. A. Chan, Patient HLA class I genotype influences cancer response to checkpoint blockade immunotherapy. Science. 359, eaao4572 (2017).

28. V. Naranbhai, M. Viard, M. Dean, S. Groha, D. A. Braun, C. Labaki, S. A. Shukla, Y. Yuki, P. Shah, K. Chin, M. Wind-Rotolo, X. J. Mu, P. B. Robbins, A. Gusev, T. K. Choueiri, J. L. Gulley, M. Carrington, HLA-A*03 and response to immune checkpoint blockade in cancer: an epidemiological biomarker study. Lancet Oncol (2021), doi:10.1016/s1470-2045(21)00582-9.

29. A. Chhibber, L. Huang, H. Zhang, J. Xu, R. Cristescu, X. Liu, D. V. Mehrotra, J. Shen, P. M. Shaw, M. D. Hellmann, A. Snyder, Germline HLA landscape does not predict efficacy of pembrolizumab monotherapy across solid tumor types. Immunity (2022), doi:10.1016/j.immuni.2021.12.006.

30. K. Litchfield, J. L. Reading, C. Puttick, K. Thakkar, C. Abbosh, R. Bentham, T. B. K. Watkins, R. Rosenthal, D. Biswas, A. Rowan, E. Lim, M. A. Bakir, V. Turati, J. A. Guerra-Assunção, L. Conde, A. J. S. Furness, S. K. Saini, S. R. Hadrup, J. Herrero, S.-H. Lee, P. V. Loo, T. Enver, J. Larkin, M. D. Hellmann, S. Turajlic, S. A. Quezada, N. McGranahan, C. Swanton, Meta-analysis of tumor- and T cell-intrinsic mechanisms of sensitization to checkpoint inhibition. Cell (2021), doi:10.1016/j.cell.2021.01.002.

31. D. Chowell, C. Krishna, F. Pierini, V. Makarov, N. A. Rizvi, F. Kuo, L. G. T. Morris, N. Riaz, T. L. Lenz, T. A. Chan, Evolutionary divergence of HLA class I genotype impacts efficacy of cancer immunotherapy. Nat Med. 25, 1715–1720 (2019).

32. M. L. Burger, A. M. Cruz, G. E. Crossland, G. Gaglia, C. C. Ritch, S. E. Blatt, A. Bhutkar, D. Canner, T. Kienka, S. Z. Tavana, A. L. Barandiaran, A. Garmilla, J. M. Schenkel, M. Hillman, I. de los R. Kobara, A. Li, A. M. Jaeger, W. L. Hwang, P. M. K. Westcott, M. P. Manos, M. M. Holovatska, F. S. Hodi, A. Regev, S. Santagata, T. Jacks, Antigen dominance hierarchies shape TCF1+ progenitor CD8 T cell phenotypes in tumors. Cell. 184, 4996-5014.e26 (2021).

33. M. Foroutan, D. D. Bhuva, R. Lyu, K. Horan, J. Cursons, M. J. Davis, Single sample scoring of molecular phenotypes. Bmc Bioinformatics. 19, 404 (2018).

34. G. A. Auwera, M. O. Carneiro, C. Hartl, R. Poplin, G. del Angel, A. Levy-Moonshine, T. Jordan, K. Shakir, D. Roazen, J. Thibault, E. Banks, K. V. Garimella, D. Altshuler, S. Gabriel, M. A. DePristo, From FastQ Data to High-Confidence Variant Calls: The Genome Analysis Toolkit Best Practices Pipeline. Curr Protoc Bioinform, in press, doi:10.1002/0471250953.bi1110s43.

35. H. Li, R. Durbin, Fast and accurate short read alignment with Burrows-Wheeler transform. Bioinform Oxf Engl. 25, 1754–60 (2009).

36. D. H. Alexander, K. Lange, Enhancements to the ADMIXTURE algorithm for individual ancestry estimation. Bmc Bioinformatics. 12, 246 (2011).

37. A. Agrawal, A. M. Chiu, M. Le, E. Halperin, S. Sankararaman, Scalable probabilistic PCA for large-scale genetic variation data. Plos Genet. 16, e1008773 (2020).

38. S. Kawaguchi, K. Higasa, M. Shimizu, R. Yamada, F. Matsuda, HLA-HD: An accurate HLA typing algorithm for next-generation sequencing data. Hum Mutat. 38, 788–797 (2017).

39. D. Roe, R. Kuang, Accurate and Efficient KIR Gene and Haplotype Inference From Genome Sequencing Reads With Novel K-mer Signatures. Front Immunol. 11, 583013 (2020).

40. M. Migdal, D. F. Ruan, W. F. Forrest, A. Horowitz, C. Hammer, MiDAS—Meaningful Immunogenetic Data at Scale. Plos Comput Biol. 17, e1009131 (2021).

41. D. Pende, M. Falco, M. Vitale, C. Cantoni, C. Vitale, E. Munari, A. Bertaina, F. Moretta, G. D. Zotto, G. Pietra, M. C. Mingari, F. Locatelli, L. Moretta, Killer Ig-Like Receptors (KIRs): Their Role in NK Cell Modulation and Developments Leading to Their Clinical Exploitation. Front Immunol. 10, 1179 (2019).

