## Supplementary Materials for "Natural killer cell educating KIR/HLA combinations impact survival in anti-PD-L1 treated cancer patients"

**This PDF file includes:**

Materials and Methods
Supplementary Text
Figs. S1 to S3
Tables S1 to S13
Appendix: List of Ethics Committees and IRBs

**Other Supplementary Materials for this manuscript include the following:**

Table S8 as separate Excel file

#### Materials and Methods

Studies and subjects

Patient data from three atezolizumab phase III clinical trials were included in this analysis: IMpower150, IMpower130, and IMpower131. Clinical trial results have been previously reported (15,17-18), and the clinical trial protocols have been provided as supplementary materials in the original study publications. Patients included in the present study signed an optional Research Biosample Repository (RBR) Informed Consent Form (ICF) and provided whole blood samples. By signing, patients provided informed consent for analysis of inherited and non-inherited genetic variation from whole blood samples. Ethics Committees (EC) and Institutional Review Boards (IRB) in each country and each study site for each clinical trial approved the clinical trial protocol, the main study ICF, and the RBR ICF. All EC and IRB forms are provided in the appendix to this supplementary document.

Whole Genome Sequencing, HLA and KIR inference

Genomic DNA was extracted from blood samples using the DNA Blood400 kit (Chemagic) and eluted in 50 μL Elution Buffer (EB, Qiagen). DNA was sheared (Covaris LE220) and sequencing libraries were prepared using the TruSeq Nano DNA HT kit (Illumina Inc.). Libraries were sequenced at Human Longevity (San Diego, CA, USA). 150bp paired-end whole-genome sequencing (WGS) data was generated to an average read depth of 30x using the HiSeq platform (Illumina X10, San Diego, CA, USA) and processed using the Burrows Wheeler Aligner (BWA)/Genome Analysis Toolkit (GATK) best practices pipeline (34). Short reads were mapped to hg38/GRCh38 (GCA_000001405.15), including alternate assemblies, using an alt-aware version of BWA to generate BAM files (35). All sequencing data was checked for concordance with data from a SNP Trace Panel (96 markers; Fluidigm, South San Francisco, CA, USA) generated before sequencing. ADMIXTURE v1.23 was used to estimate ancestry in the 5 major populations using supervised mode (36). Only samples with >0.8 European (EUR) ancestry were included in our analyses. Genetic principal component analysis (PCA) was performed using the ProPCA algorithm (37).

We used HLA-HD to infer HLA alleles from whole-genome sequencing data at 3-field resolution (38). KIR genotyping was performed with KPI v1.0.1 (39), starting from BAM files generated as described above. The MiDAS package v1.1.0 for R was used to infer KIR-HLA combinations from the HLA allele and KIR presence/absence calls, HLA class I supertypes, as well as for analyses of HLA heterozygosity and evolutionary divergence (40).

IMpower150 RNA-sequencing data

Formalin-fixed paraffin-embedded tissue (FFPET) was macro-dissected for the tumor area. RNA was extracted using the High Pure FFPET RNA Isolation Kit (Roche) and assessed by Qubit and Agilent Bioanalyzer for quantity and quality. First strand cDNA synthesis was performed from total RNA using random primers, followed by the generation of second strand cDNA with dUTP in place of dTTP in the master mix to facilitate preservation of strand information. Libraries were enriched for the mRNA fraction by positive selection using a cocktail of biotinylated oligos corresponding to coding regions of the genome. Libraries were sequenced using Illumina HiSeq.

TruSeq technology (Illumina) was used to generate whole-transcriptome profiles. To remove ribosomal reads, RNA-seq reads were first aligned to ribosomal RNA sequences. GSNAP version

2013-10-10 was used to align the remaining reads to the human reference genome (NCBI Build 38), allowing a maximum of two mismatches per 75 base sequences (parameters: '-M 2 -n 10 -B 2 -i 1 -N 1 -w 200000 -E 1-pairmax-rna = 200000 –clip-overlap). To quantify gene expression levels, the number of reads mapped to the exons of each RefSeq gene was calculated using the functionality provided by the R/Bioconductor package GenomicAlignments.

An NK infiltration score was implemented as described by Cursons et al. (4). In short, a gene set was derived that signaled abundance of NK cells. The original list of candidate genes was taken from previously-published human and mouse expression profiles. This candidate list was filtered based on solid tumor expression levels from several other previously-published studies. The resulting set of 20 genes (*CD160, CD244, CTSW, FASLG, GZMA, GZMB, GZMH, IL18RAP, IL2RB, KIR2DL4, KLRB1, KLRC3, KLRD1, KLRF1, KLRK1, NCR1, NKG7, PRF1, XCL1, XCL2*) was used with the singscore rank-based algorithm to estimate NK cell infiltration (33).

<u>Statistical analysis</u>

Cox proportional hazards models were used to test for association of all tested genetic variables (HLA alleles, HLA ligand groups, KIR genes, KIR-HLA combinations) and the NK cell score with overall survival. Due to the complex linkage disequilibrium among HLA ligand groups and KIR gene copy number, it is difficult to assume statistical independence. Thus, statistics for each genetic variable were derived from multivariable models incorporating all tested KIR/HLA interactions, HLA ligand groups, or KIR genes. Age, sex, ECOG performance status, and the first three genetic principal components (PCs) were included as covariates. For the replication cohort 1 from Chowell et al., we included age, gender and drug class as covariates, and for the MSK-IMPACT replication cohort age group and drug class (27). To assess pairwise differences between >2 survival curves, the "pairwise_survdiff" function from the R package survminer was applied, and resulting p-values are Benjamini-Hochberg adjusted.

For HLA allelic association analyses (2-field resolution and supertypes), a dominant inheritance model was chosen (carrier vs. non-carrier), and only alleles with a carrier frequency of >0.5% in each study were included in the analyses.

For the meta-analyses, the random-effects model method of the metagen function in the R package "meta" was used to calculate effect estimates, 95% confidence intervals, p-values, and between-study variance using the DerSimonian-Laird method. No significant heterogeneity between studies was identified for any of the performed meta-analyses.

The Bonferroni method was applied to correct P values for multiple testing, and P values are reported uncorrected, unless specified otherwise.

Linear regression was performed to understand the relationship between KIR/HLA combinations and the NK cell signature, using age, sex, ECOG performance status, and three PCs.

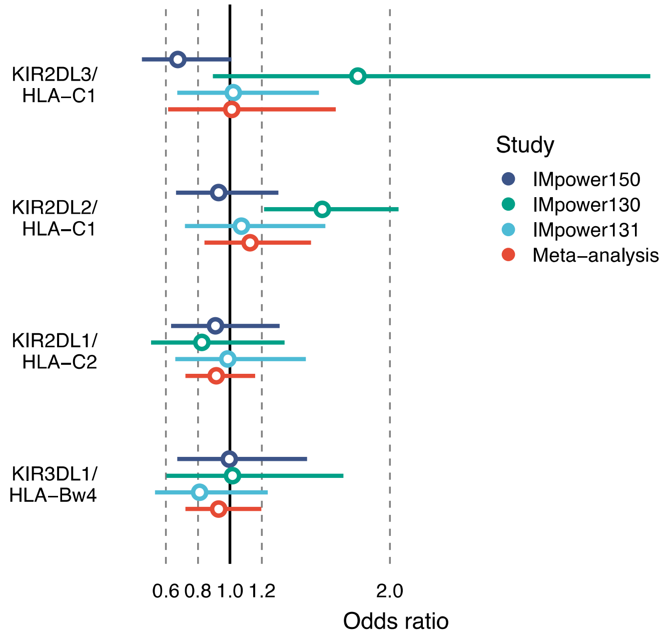


**Fig. S1.**

Association between KIR/HLA combinations and overall survival for patients with NSCLC in the control arms of IMpower150 (N=182), IMpower130 (N=102) and IMpower131 (N=156), as well as meta-analysis results (N=440). Horizontal lines depict 95% confidence intervals. The complete data are provided in Table S3.

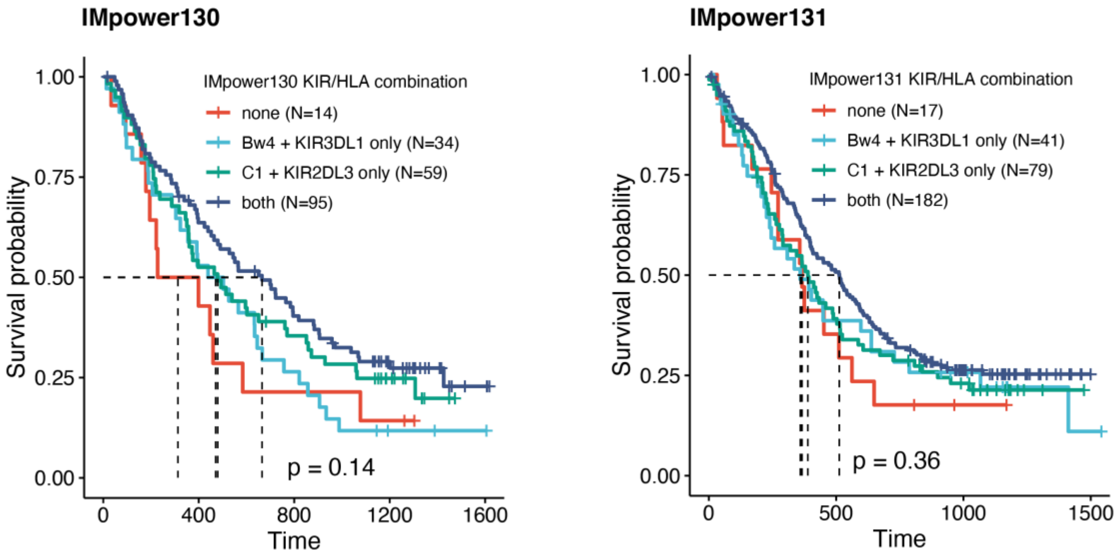


**Fig. S2.**

Kaplan-Meier estimates of overall survival in IMpower130 and IMpower131 for atezolizumab-treated patients carrying none of the two significant KIR/HLA interactions KIR3DL1-Bw4 and KIR2DL3/HLA-C1 (N=20), one of the two (N=81 and N=114, respectively), and both (N=218). Dashed lines depict median survival for each group. P-value is based on a log-rank test.

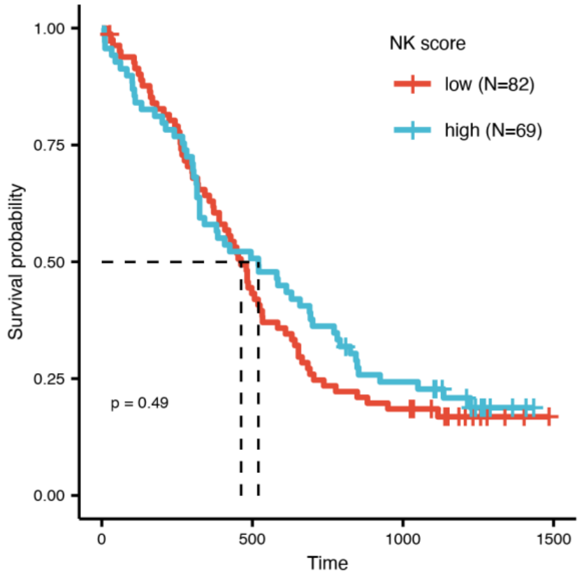


**Fig. S3.**

Kaplan-Meier estimates of overall survival for patients in IMpower150 control arms (no immunotherapy) with low (below median, N=82) and high (above median, N=69) NK cell score. Dashed lines depict median survival for each group. P-value is based on a log-rank test.

**Table S1.**

List of included clinical trials with description of experimental and comparator arms.

| Study | Clinicaltrials.gov ID | Cancer type | Study arms |
|---|---|---|---|
| **IMpower150** | NCT02366143 | Stage IV Non-Squamous Non-Small Cell Lung Cancer | Experimental: Arm A (Atezolizumab+Paclitaxel+Carboplatin) |
| | | | Experimental: Arm B (Atezolizumab+Bevacizumab+Paclitaxel + Carboplatin) |
| | | | Active Comparator: Arm C (Bevacizumab+Paclitaxel+Carboplatin) |
| **IMpower130** | NCT02367781 | Stage IV Non-Squamous Non-Small Cell Lung Cancer | Experimental: Arm A (Atezolizumab+Nab-Paclitaxel+Carboplatin) |
| | | | Active Comparator: Arm B (Nab-Paclitaxel+Carboplatin) |
| **IMpower131** | NCT02367794 | Stage IV Squamous Non-Small Cell Lung Cancer | Experimental: Arm A: Atezolizumab + Paclitaxel + Carboplatin |
| | | | Experimental: Arm B: Atezolizumab + Nab-Paclitaxel + Carboplatin |
| | | | Active Comparator: Arm C: Nab-Paclitaxel + Carboplatin |

**Table S2.**

Carrier frequencies for HLA ligands, KIR genes, and KIR/HLA combinations in the included IMpower trials, as well as published reference frequencies.

| | Carrier frequency | | | | |
|---|---|---|---|---|---|
| **Variable** | **IMpower150** | **IMpower130** | **IMpower131** | **EUR Ref min** | **EUR Ref max** |
| **HLA-C1** | 0.846 | 0.891 | 0.857 | | |
| **HLA-C2** | 0.641 | 0.599 | 0.6 | | |
| **HLA-Bw4** | 0.745 | 0.651 | 0.743 | | |
| **KIR2DL1** | 0.98 | 0.967 | 0.987 | 0.878 | 1 |
| **KIR2DL2** | 0.541 | 0.493 | 0.533 | 0.392 | 0.656 |
| **KIR2DL3** | 0.901 | 0.891 | 0.916 | 0.565 | 0.961 |
| **KIR3DL1** | 0.959 | 0.954 | 0.945 | 0.758 | 1 |
| **KIR2DL2/HLA-C1** | 0.46 | 0.441 | 0.457 | 0.316 | 0.52 |
| **KIR2DL3/HLA-C1** | 0.763 | 0.789 | 0.792 | 0.526 | 0.824 |
| **KIR2DL1/HLA-C2** | 0.633 | 0.586 | 0.596 | 0.396 | 0.677 |
| **KIR3DL1/HLA-Bw4** | 0.709 | 0.625 | 0.701 | 0.283 | 0.664 |

KIR reference data was retrieved from http://www.allelefrequencies.net/kir.asp, filtering for Caucasoid ethnicity, N>100, and Resolution=0
KIR/HLA reference data was retrieved from http://www.allelefrequencies.net/KIR-HLA/stats.asp

**Table S3.**

Association analysis results for KIR/HLA combinations and overall survival in IMpower150

| | Atezolizumab (N=433) | | | Control arm (N=181) | | |
|---|---|---|---|---|---|---|
| **HLA-KIR interaction** | **HR** | **95% CI** | **P value** | **HR** | **95% CI** | **P value** |
| **KIR2DL3/HLA-C1** | **0.62** | **0.47-0.80** | **3.4E-04** | 0.67 | 0.45-1.01 | 5.5E-02 |
| **KIR2DL2/HLA-C1** | 1.12 | 0.90-1.41 | 3.1E-01 | 0.93 | 0.66-1.30 | 6.7E-01 |
| **KIR2DL1/HLA-C2** | 0.80 | 0.63-1.02 | 7.3E-02 | 0.91 | 0.63-1.31 | 6.1E-01 |
| **KIR3DL1/HLA-Bw4** | 0.88 | 0.68-1.13 | 3.2E-01 | 1.00 | 0.67-1.48 | 9.9E-01 |

**Table S4.**

Association results for KIR/HLA combinations and overall survival in the IMpower trials and meta-analysis

| Study | KIR/HLA combination | Atezolizumab | | | | Control arms | | | |
|---|---|---|---|---|---|---|---|---|---|
| | | N | HR | 95% CI | P value | N | HR | 95% CI | P value |
| **IMpower150** | KIR2DL3/HLA-C1 | 433 | **0.62** | **0.47-0.80** | **3.4E-04** | 182 | 0.67 | 0.45-1.01 | 5.5E-02 |
| | KIR2DL2/HLA-C1 | 433 | 1.12 | 0.90-1.41 | 3.1E-01 | 182 | 0.93 | 0.66-1.30 | 6.7E-01 |
| | KIR2DL1/HLA-C2 | 433 | 0.80 | 0.63-1.02 | 7.3E-02 | 182 | 0.91 | 0.63-1.31 | 6.1E-01 |
| | KIR3DL1/HLA-Bw4 | 433 | 0.88 | 0.68-1.13 | 3.2E-01 | 182 | 1.00 | 0.67-1.48 | 9.9E-01 |
| **IMpower130** | KIR2DL3/HLA-C1 | 202 | *0.69* | *0.48-0.99* | *4.4E-02* | 102 | 1.80 | 0.89-3.63 | 1.0E-01 |
| | KIR2DL2/HLA-C1 | 202 | 1.05 | 0.75-1.47 | 7.7E-01 | 102 | 1.58 | 1.00-2.50 | 5.2E-02 |
| | KIR2DL1/HLA-C2 | 202 | 1.06 | 0.74-1.50 | 7.5E-01 | 102 | 0.82 | 0.51-1.34 | 4.4E-01 |
| | KIR3DL1/HLA-Bw4 | 202 | 0.72 | 0.50-1.03 | 7.4E-02 | 102 | 1.02 | 0.60-1.71 | 9.6E-01 |
| **IMpower131** | KIR2DL3/HLA-C1 | 319 | 0.82 | 0.58-1.16 | 2.6E-01 | 156 | 1.02 | 0.67-1.56 | 9.2E-01 |
| | KIR2DL2/HLA-C1 | 319 | 1.04 | 0.80-1.35 | 7.8E-01 | 156 | 1.07 | 0.72-1.60 | 7.3E-01 |
| | KIR2DL1/HLA-C2 | 319 | 1.04 | 0.78-1.37 | 8.1E-01 | 156 | 0.99 | 0.66-1.47 | 9.4E-01 |
| | KIR3DL1/HLA-Bw4 | 319 | 0.82 | 0.62-1.10 | 1.8E-01 | 156 | 0.81 | 0.53-1.24 | 3.3E-01 |
| **Meta-analysis** | KIR2DL3/HLA-C1 | 954 | **0.68** | **0.57-0.82** | **5.0E-05** | 440 | 1.01 | 0.60-1.68 | 9.7E-01 |
| | KIR2DL2/HLA-C1 | 954 | 1.08 | 0.92-1.26 | 3.2E-01 | 440 | 1.13 | 0.84-1.51 | 4.3E-01 |
| | KIR2DL1/HLA-C2 | 954 | 0.93 | 0.77-1.13 | 4.9E-01 | 440 | 0.91 | 0.72-1.16 | 4.6E-01 |
| | KIR3DL1/HLA-Bw4 | 954 | *0.82* | *0.70-0.97* | *2.4E-02* | 440 | 0.93 | 0.72-1.20 | 5.7E-01 |

**Table S5.**

Pairwise comparisons of survival curves for groups according to KIR/HLA combination carrier status. P-values are Benjamini-Hochberg adjusted and based on log-rank tests.

| Study | Interactions | none | KIR3DL1/HLA-Bw4 only | KIR2DL3/HLA-C1 only |
|---|---|---|---|---|
| IMpower150 | KIR3DL1/HLA-Bw4 only | 0.370 | | 0.250 |
| | KIR2DL3/HLA-C1 only | 0.188 | 0.250 | |
| | Both | **0.037** | **0.019** | 0.250 |
| IMpower130 | KIR3DL1/HLA-Bw4 only | 0.770 | | 0.420 |
| | KIR2DL3/HLA-C1 only | 0.420 | 0.420 | |
| | both | 0.290 | 0.250 | 0.440 |
| IMpower131 | KIR3DL1/HLA-Bw4 only | 0.860 | | 0.920 |
| | KIR2DL3/HLA-C1 only | 0.860 | 0.920 | |
| | both | 0.560 | 0.560 | 0.560 |

**Table S6.**

Association results for HLA ligand carrier status and overall survival in the IMpower trials and replication trials from Chowell et al., as well as meta-analysis

| Study | HLA group | HR | 95% CI | P value |
|---|---|---|---|---|
| **IMpower150** | HLA-C1 | 0.79 | 0.59-1.07 | 1.3E-01 |
| | HLA-Bw4 | *0.77* | *0.60-0.99* | *3.9E-02* |
| **IMpower130** | HLA-C1 | *0.59* | *0.37-0.94* | *2.6E-02* |
| | HLA-Bw4 | *0.67* | *0.47-0.96* | *2.9E-02* |
| **IMpower131** | HLA-C1 | 0.69 | 0.48-1.01 | 5.6E-02 |
| | HLA-Bw4 | 0.80 | 0.60-1.06 | 1.2E-01 |
| **Chowell et al.** | HLA-C1 | 0.63 | 0.31-1.30 | 1.3E-01 |
| | HLA-Bw4 | 0.84 | 0.41-1.73 | 1.7E-01 |
| **MSK-IMPACT** | HLA-C1 | 0.75 | 0.43-1.33 | 3.2E-01 |
| | HLA-Bw4 | 0.91 | 0.58-1.44 | 6.9E-01 |
| **Meta analysis** | HLA-C1 | **0.71** | **0.59-0.86** | **4.9E-04** |
| | HLA-Bw4 | **0.77** | **0.66-0.90** | **1.1E-03** |

**Table S7.**

Association results for KIR gene presence and overall survival in the IMpower and meta-analysis

| Study | KIR gene | HR | 95% CI | P value |
|---|---|---|---|---|
| **IMpower150** | *KIR2DL3* | **0.56** | **0.39-0.78** | **8.2E-04** |
| | *KIR3DL1* | 1.69 | 0.90-3.18 | 1.0E-01 |
| **IMpower130** | *KIR2DL3* | 0.83 | 0.51-1.36 | 4.6E-01 |
| | *KIR3DL1* | 1.26 | 0.50-3.18 | 6.2E-01 |
| **IMpower131** | *KIR2DL3* | 1.18 | 0.71-1.95 | 5.2E-01 |
| | *KIR3DL1* | 1.25 | 0.66-2.38 | 5.0E-01 |
| **Meta analysis** | *KIR2DL3* | 0.79 | 0.51-1.23 | 3.0E-01 |
| | *KIR3DL1* | 1.42 | 0.95-2.13 | 9.2E-02 |

**Table S8.**

HLA class I and class II single-allele associations with overall survival in NSCLC patients treated with atezolizumab, based on a meta-analysis of the IMpower150, IMpower130, and IMpower131 trials.

Table is provided in Other Supplementary Material as an Excel file.

**Table S9.**

HLA class I supertype associations with overall survival in NSCLC patients treated with atezolizumab, based on a meta-analysis of the IMpower150, IMpower130, and IMpower131 trials.

| HLA supertype | P value | P value (adj) | HR | SE |
|---|---|---|---|---|
| **A01** | 0.255 | 1 | 1.090 | 0.076 |
| **A01A03** | 0.638 | 1 | 1.219 | 0.421 |
| **A01A24** | 0.730 | 1 | 0.945 | 0.163 |
| **A02** | 0.157 | 1 | 0.899 | 0.076 |
| **A03** | 0.787 | 1 | 1.034 | 0.125 |
| **A24** | 0.360 | 1 | 0.918 | 0.093 |
| **B07** | 0.534 | 1 | 1.048 | 0.076 |
| **B08** | 0.535 | 1 | 1.112 | 0.172 |
| **B27** | 0.890 | 1 | 0.988 | 0.091 |
| **B44** | 0.986 | 1 | 0.999 | 0.076 |
| **B58** | 0.420 | 1 | 0.851 | 0.200 |
| **B62** | 0.555 | 1 | 0.932 | 0.120 |

**Table S10.**

HLA class I and class II heterozygosity associations with overall survival in NSCLC patients treated with atezolizumab, based on a meta-analysis of the IMpower150, IMpower130, and IMpower131 trials.

| HLA gene | P value | P value (adj) | HR | SE |
|---|---|---|---|---|
| ***A*** | 0.681 | 1 | 1.046 | 0.108 |
| ***B*** | 0.124 | 1 | 0.808 | 0.139 |
| ***C*** | 0.164 | 1 | 0.849 | 0.117 |
| ***DPA1*** | 0.882 | 1 | 0.988 | 0.082 |
| ***DPB1*** | 0.451 | 1 | 0.926 | 0.103 |
| ***DQA1*** | 0.334 | 1 | 0.901 | 0.108 |
| ***DQB1*** | 0.545 | 1 | 0.929 | 0.121 |
| ***DRA*** | 0.943 | 1 | 1.007 | 0.093 |
| ***DRB1*** | 0.227 | 1 | 0.861 | 0.124 |

**Table S11.**

HLA class I evolutionary divergence (Grantham distance) associations with overall survival in NSCLC patients treated with atezolizumab, based on a meta-analysis of the IMpower150, IMpower130, and IMpower131 trials.

| HLA gene | P value | P value (adj) | HR | SE |
|---|---|---|---|---|
| ***A*** | 0.959 | 1.000 | 1.001 | 0.010 |
| ***B*** | 0.096 | 0.385 | 0.970 | 0.018 |
| ***C*** | *0.036* | *0.143* | *0.977* | *0.011* |
| **ABC_average** | 0.141 | 0.564 | 0.975 | 0.017 |

**Table S12.**

Multivariable analysis results for NK cell score (below vs. above median) and KIR/HLA combinations, and overall survival in atezolizumab-treated patients in IMpower150.

| | IMpower150 atezolizumab arms (N=371) | | |
|---|---|---|---|
| **Variable** | **HR** | **95% CI** | **P value** |
| NK cell score > median | **0.74** | **0.58-0.94** | **0.015** |
| KIR2DL3/HLA-C1 | **0.69** | **0.51-0.92** | **0.012** |
| KIR2DL2/HLA-C1 | 1.07 | 0.84-1.36 | 0.569 |
| KIR2DL1/HLA-C2 | 0.83 | 0.64-1.07 | 0.153 |
| KIR3DL1/HLA-Bw4 | 0.96 | 0.73-1.26 | 0.759 |

**Table S13.**

Pairwise comparisons of survival curves for groups according to NK score and KIR2DL3/HLA-C1 carrier status in IMpower150 (atezolizumab-treated patients). P-values are Benjamini-Hochberg adjusted and based on log-rank tests.

| **Interactions** | **NK score low, no KIR2DL3/HLA-C1** | **NK score high, no KIR2DL3/HLA-C1** | **NK score low, KIR2DL3/HLA-C1 carrier** |
|---|---|---|---|
| NK score high, no KIR2DL3/HLA-C1 | 0.174 | | |
| NK score low, KIR2DL3/HLA-C1 carrier | 0.156 | 0.968 | |
| NK score high, KIR2DL3/HLA-C1 carrier | **0.008** | 0.169 | 0.076 |

#### Appendix

List of Ethics Committees and Institutional Review Boards (IRB) for IMpower150, IMpower130, and IMpower131

### IMpower150

#### Names and Addresses of Institutional Review Boards / Ethics Committees
#### Protocol: GO29436

| Site # | Investigator | IRB/EC Name and Address | Approval Date |
|---|---|---|---|
| 278207 | Telivala, Bijoy | Copernicus Group Independent Review Board; 1 Triangle Drive Suite 100 PO Box 110605 Research Triangle Park North Carolina 27709 United States | 8-Apr-15 |
| 278208 | Hamm, John | Western Institutional Review Board; 1019 39th Avenue Southeast Suite 120 Puyallup Washington 98374 United States | 3-Jun-15 |
| 278209 | Shtivelband, Mikhael | Copernicus Group Independent Review Board; 1 Triangle Drive Suite 100 PO Box 110605 Research Triangle Park North Carolina 27709 United States | 8-Jun-15 |
| 278210 | Erickson, Brian | Copernicus Group Independent Review Board; 1 Triangle Drive Suite 100 PO Box 110605 Research Triangle Park North Carolina 27709 United States | 25-Feb-15 |
| 278223 | John, Thomas | Melbourne Health Human Research Ethics Committee | 4-Dec-15 |
| 278225 | Hughes, Brett | Melbourne Health Human Research Ethics Committee | 21-Apr-15 |
| 278265 | Crombie, Catherine | Melbourne Health Human Research Ethics Committee | 21-Apr-15 |
| 278283 | Hooberman, Arthur | Copernicus Group Independent Review Board; 1 Triangle Drive Suite 100 PO Box 110605 Research Triangle Park North Carolina 27709 United States | 4-Mar-15 |
| 278738 | Blinman, Prunella | Melbourne Health Human Research Ethics Committee | 21-Apr-15 |
| 278740 | Gauden, Stan | Tasmania Health and Medical Human Research Ethics Committee | 30-Mar-15 |
| 278746 | Potasz, Nicole | Melbourne Health Human Research Ethics Committee | 21-Apr-15 |
| 278748 | Singhal, Nimit | Melbourne Health Human Research Ethics Committee | 21-Apr-15 |
| 278754 | O'Byrne, Kenneth | Melbourne Health Human Research Ethics Committee | 21-Apr-15 |
| 278824 | Aleman Polanco, Diana Sofía | Asociacion Benefica Prisma | 23-Apr-15 |
| 278825 | Mas Lopez, Luis | Comité Institucional de ética en Investigación Instituto Nacional de Enfermedades Neoplasicas | 4-May-15 |
| 278827 | Lim, Darren | Singhealth Centralised Institutional Review Board | 10-Mar-15 |
| 278837 | Hung, Jen-Yu | Institutional Review Board Kaohsiung Medical University Chung-Ho Memorial Hospital | 30-May-15 |
| 278841 | Chen, Yuh-Min | Institutional Review Board Taipei Veterans General Hospital | 30-Mar-16 |
| 278844 | Li, Chien-Te | Institutional Review Board, Changhua Christian Hospital | 9-Jun-15 |
| 278847 | Chen, Chao-Hsun | Institutional Review Board of the Chi Mei Medical Center | 2-Jun-15 |
| 278849 | Chang, Gee-Chen | The Institutional Review Board of Taichung Veterans General Hospital | 8-Jun-15 |
| 278850 | Ho, Ching-Liang | Institutional Review Board of Tri-Service General Hospital | 13-Jun-15 |
| 279286 | Chen, Wei-Teing | Cheng-Hsin General Hospital Institutional Review Board | 9-Jul-15 |
| 279291 | Magri, Ignacio | Consejo de Evaluación Ética de Investigación en Salud - CoEIS | 14-Jul-16 |
| 279293 | Kowalyszyn, Rubén | Ministerio de Salud - Provincia de Río Negro | 12-Nov-15 |
| 279293 | Kowalyszyn, Rubén | Comité Independiente de Etica en investigación clínica "Dr. Carlos A. Barclay | 12-Nov-15 |
| 279295 | Lerzo, Guillermo | Comité Independiente de Etica en investigación clínica "Dr. Carlos A. Barclay | 12-Feb-15 |
| 279300 | Jarchum, Gustavo | Consejo de Evaluación Ética de Investigación en Salud - CoEIS | 1-Dec-15 |
| 279302 | Kaen, Diego | Comité Independiente de Etica en investigación clínica "Dr. Carlos A. Barclay | 19-Feb-15 |
| 279305 | Kahl, Susana | Comité de ética en investigación Fundación Oncosalud | 20-Feb-16 |
| 279305 | Kahl, Susana | Comisión Conjunta de Investigación en Salud (CCIS) | 20-Feb-16 |

| Site # | Investigator | IRB/EC Name and Address | Approval Date |
|---|---|---|---|
| 279308 | Varela, Mirta | Comité de Ética Centro de Oncología e Investigación Buenos Aires | 6-May-15 |
| 279308 | Varela, Mirta | Comisión Conjunta de Investigación en Salud (CCIS) | 6-May-15 |
| 279311 | Kotliar, Mauricio | Comité de Ética Independiente - Fundación Sanatorio | 28-Apr-15 |
| 279317 | Lin, Yu-Ching | Institutional Review Board of Chang Gung Medical Foundation | 11-Jun-15 |
| 279318 | Eckmayr, Josef | Ethikkommission für das Bundesland Salzburg; Sebastian-Stief-Gasse 2 Salzburg Salzburg 5010 Austria | 16-Oct-15 |
| 279322 | Zvirbule, Zanete | The Ethics Committee for Clinical Trials of Medicinal Products | 24-Apr-15 |
| 279787 | Drew, David | Copernicus Group Independent Review Board; 1 Triangle Drive Suite 100 PO Box 110605 Research Triangle Park North Carolina 27709 United States | 1-Apr-15 |
| 279788 | Almubarak, Mohammed | Chesapeake Institutional Review Board; 6940 Columbia Gateway Drive Suite 110 Columbia Maryland 21046 United States | 13-ago-15 |
| 279789 | Assikis, Vasileios | Copernicus Group Independent Review Board; 1 Triangle Drive Suite 100 PO Box 110605 Research Triangle Park North Carolina 27709 United States | 9-Apr-15 |
| 279819 | Belman, Neil | St. Luke’s Hospital & Health Network IRB; 801 Ostrum Street East Wing 4 Bethlehem Pennsylvania 18015 United States | 25-Mar-15 |
| 279820 | Chaudhry, Arvind | Copernicus Group Independent Review Board; 1 Triangle Drive Suite 100 PO Box 110605 Research Triangle Park North Carolina 27709 United States | 3-Feb-15 |
| 279821 | Chitneni, Shobha | Copernicus Group Independent Review Board; 1 Triangle Drive Suite 100 PO Box 110605 Research Triangle Park North Carolina 27709 United States | 29-Jan-15 |
| 279822 | Cohenuram, Michael | Copernicus Group Independent Review Board; 1 Triangle Drive Suite 100 PO Box 110605 Research Triangle Park North Carolina 27709 United States | 13-Mar-15 |
| 279823 | De Vore, Russell | Copernicus Group Independent Review Board; 1 Triangle Drive Suite 100 PO Box 110605 Research Triangle Park North Carolina 27709 United States | 3-Jun-15 |
| 279824 | Goldschmidt, Jerome | Copernicus Group Independent Review Board; 1 Triangle Drive Suite 100 PO Box 110605 Research Triangle Park North Carolina 27709 United States | 4-Feb-15 |
| 279825 | Halibey, Bohdan | Copernicus Group Independent Review Board; 1 Triangle Drive Suite 100 PO Box 110605 Research Triangle Park North Carolina 27709 United States | 10-Apr-15 |
| 279826 | Kirshner, Eli | Western Institutional Review Board; 1019 39th Avenue Southeast Suite 120 Puyallup Washington 98374 United States | 31-mar-15 |
| 279827 | Martin, William | St Charles Medical Center; 2500 Northeast Neff Road Bend Oregon 97701 United States | 25-mar-15 |
| 279828 | Mitchell, Reed | Copernicus Group Independent Review Board; 1 Triangle Drive Suite 100 PO Box 110605 Research Triangle Park North Carolina 27709 United States | 26-Jan-15 |
| 279829 | Subramanian, Janakiraman | Saint Luke's Hospital Institutional Review Board; 915 East First Street Duluth Minnesota 55805 United States | 8-Jun-15 |
| 279830 | Swanson, Paul | Copernicus Group Independent Review Board; 1 Triangle Drive Suite 100 PO Box 110605 Research Triangle Park North Carolina 27709 United States | 20-Feb-15 |
| 279831 | Tsai, Frank Yung-Chin | Western Institutional Review Board; 1019 39th Avenue Southeast Suite 120 Puyallup Washington 98374 United States | 8-Jul-15 |
| 279832 | Abdel Karim, Nagla | Western Institutional Review Board; 1019 39th Avenue Southeast Suite 120 Puyallup Washington 98374 United States | 19-feb-16 |
| 279833 | Früh, Martin | Kantonale Ethikkommission Bern (KEK) | 3-Mar-16 |
| 279836 | Orlandi Jorquera, Francisco | Comite de Etica Cientifico del Servicio de Salud Metropolitano Norte | 26-May-15 |
| 279838 | Allegrini, Giacomo | Comitato Etico Area Vasta Nord Ovest presso Azienda Ospedaliero Universitaria Pisana di Pisa | 9-Jul-15 |

| Site # | Investigator | IRB/EC Name and Address | Approval Date |
|---|---|---|---|
| 279839 | Cartenì, Giacomo | Comitato Etico Cardarelli-Santobono | 7-Oct-15 |
| 279840 | Falcone, Alfredo | Comitato Etico Area Vasta Nord Ovest presso Azienda Ospedaliero Universitaria Pisana di Pisa | 9-Jul-15 |
| 279841 | Mencoboni, Manlio | Comitato Etico San Martino IST 2 | 16-Jul-15 |
| 279843 | Migliorino, Maria | Comitato Etico Lazio 1 | 30-Jul-15 |
| 279844 | Tonini, Giuseppe | COMITATO ETICO DELL´UNIVERSITA´ CAMPUS BIO-MEDICO DI ROMA | 18-Jun-15 |
| 279845 | Amoroso, Domenico | Comitato Etico Area Vasta Nord Ovest presso Azienda Ospedaliero Universitaria Pisana di Pisa | 9-Jul-15 |
| 279846 | Biesma, Bonne | Medical Research Ethics Committees United | 29-Oct-15 |
| 279847 | Wilschut, Frank | Medical Research Ethics Committees United | 29-Oct-15 |
| 279848 | Moro-Sibilot, Denis | CPP Sud-Méditerranée 2; 270 Boulevard Sainte Marguerite Hôpital Sainte Margeurite Pavillon 9 Cedex 9 Marseille Bouches-du-Rhône 13274 France | 5-Jun-15 |
| 279849 | Kosmider, Suzanne | Melbourne Health Human Research Ethics Committee | 21-Apr-15 |
| 279850 | Millward, Michael | Sir Charles Gairdner Hospital HREC | 28-Jul-15 |
| 279851 | Parnis, Francis | Bellberry Human Research Ethics Committee | 5-May-15 |
| 279852 | Lewis, Craig | Melbourne Health Human Research Ethics Committee | 21-Apr-15 |
| 280022 | Richardson, Gary | Cabrini Human Research Ethics Committee | 29-Jul-15 |
| 280023 | Gill, Sanjeev | Melbourne Health Human Research Ethics Committee | 20-May-15 |
| 280024 | Soto-Parra, Hector | Comitato Etico Catania 1 presso A.O. Universitaria Policlinico Vittorio Emanuele di Catania | 9-Jun-15 |
| 280029 | Almodovar, Maria Teresa | Comissão de Ética para a Investigação Clínica - CEIC | 10-Jul-15 |
| 280031 | Araújo, António | Comissão de Ética para a Investigação Clínica - CEIC | 10-Jul-15 |
| 280032 | Barata, Fernando | Comissão de Ética para a Investigação Clínica - CEIC | 10-Jul-15 |
| 280033 | Queiroga, Henrique | Comissão de Ética para a Investigação Clínica - CEIC | 10-Jul-15 |
| 280034 | Azevedo, Isabel | Comissão de Ética para a Investigação Clínica - CEIC | 10-Jul-15 |
| 280035 | Wesseler, Claas | Ethikkommission an der Universität Regensburg; Landshuter Straße 4 Raum 134, 1.OG. Regensburg 93047 Germany | 3-Nov-15 |
| 280037 | Wermke, Martin | Ethikkommission an der Universität Regensburg | 3-Nov-15 |
| 280038 | Wehler, Thomas | Ethikkommission an der Universität Regensburg; Landshuter Straße 4 Raum 134, 1.OG. Regensburg 93047 Germany | 3-Nov-15 |
| 280039 | Loges, Sonja | Ethikkommission an der Universität Regensburg | 3-Nov-15 |
| 280040 | Serke, Monika | Ethikkommission an der Universität Regensburg; Landshuter Straße 4 Raum 134, 1.OG. Regensburg 93047 Germany | 3-Nov-15 |
| 280043 | Kokowski, Konrad | Ethikkommission an der Universität Regensburg; Landshuter Straße 4 Raum 134, 1.OG. Regensburg 93047 Germany | 3-Nov-15 |
| 280044 | Schulz, Christian | Ethikkommission an der Universität Regensburg; Landshuter Straße 4 Raum 134, 1.OG. Regensburg 93047 Germany | 3-Nov-15 |
| 280046 | Fischer, Jürgen | Ethikkommission an der Universität Regensburg; Landshuter Straße 4 Raum 134, 1.OG. Regensburg 93047 Germany | 3-Nov-15 |
| 280048 | Behringer, Dirk | Ethikkommission an der Universität Regensburg; Landshuter Straße 4 Raum 134, 1.OG. Regensburg 93047 Germany | 3-Nov-15 |
| 280049 | Bargon, Joachim | Ethikkommission an der Universität Regensburg; Landshuter Straße 4 Raum 134, 1.OG. Regensburg 93047 Germany | 3-Nov-15 |
| 280052 | Rothenstein, Jeffrey | Lakeridge Health REB; 1 Hospital Court Oshawa Ontario L1G 2B9 Canada | 15-Jul-15 |

| Site # | Investigator | IRB/EC Name and Address | Approval Date |
|---|---|---|---|
| 280255 | Dobrescu, Andrei | Copernicus Group Independent Review Board; 1 Triangle Drive Suite 100 PO Box 110605 Research Triangle Park North Carolina 27709 United States | 8-May-15 |
| 280256 | Stella, Philip | St. Joseph Mercy Health System Institutional Review Board #2 - Oncology Central IRB; 5301 East Huron River Drive Clinical Research Department, RHB 6017 Ann Arbor Michigan 48106 United States | 16-jul-15 |
| 280258 | Alnsour, Mohammad | Mercy Saint Vincent Medical Center Institutional Review Board; 2213 Cherry Street Toledo Ohio 43608 United States | 13-Apr-15 |
| 280259 | Lukas, Jason | Western Institutional Review Board; 1019 39th Avenue Southeast Suite 120 Puyallup Washington 98374 United States | 3-Jun-15 |
| 280260 | Jotte, Robert | US Oncology Inc. Institutional Review Board; 10101 Woodloch Forest The Woodlands Texas 77380 United States | 18-Jun-15 |
| 280262 | Yanagihara, Ronald | Western Institutional Review Board (WIRB); 1019 39th Avenue Southeast Suite 120 Puyallup Washington 98374 United States | 26-Jan-16 |
| 280263 | Finley, Gene | Copernicus Group Independent Review Board; 1 Triangle Drive Suite 100 PO Box 110605 Research Triangle Park North Carolina 27709 United States | 16-Jun-15 |
| 280264 | Thomas, Christian | Copernicus Group Independent Review Board; 1 Triangle Drive Suite 100 PO Box 110605 Research Triangle Park North Carolina 27709 United States | 11-May-15 |
| 280265 | Silberberg, Jeffrey | Copernicus Group Independent Review Board; 1 Triangle Drive Suite 100 PO Box 110605 Research Triangle Park North Carolina 27709 United States | 8-May-15 |
| 280267 | Garcia, Yolanda | CEIC de la Corporacion Sanitaria del Parc Tauli | 9-Jun-15 |
| 280268 | Lopez Brea, Marta | CEIC de Cantabria | 9-Jun-15 |
| 280269 | Barneto-Aranda, Isidoro | CEIC de Andalucia (CCEIBA) | 9-Jun-15 |
| 280270 | De Castro Carpeño, Javier | CEIC Hospital Universitario La Paz | 9-Jun-15 |
| 280271 | Domine Gomez, Manuel | CEIC Fundación Jiménez Díaz | 9-Jun-15 |
| 280272 | Insa Molla, Amelia | CEIC Hospital Clínico Universitario de Valencia | 9-Jun-15 |
| 280273 | Alvarez, Rosa | CEIC Hospital General Universitario Gregorio Marañon | 9-Jun-15 |
| 280274 | Gonzalez Larriba, Jose Luis | CEIC Hospital Clinico San Carlos | 9-Jun-15 |
| 280275 | Jiménez Munarriz, Beatriz | CEIC Grupo Hospital de Madrid | 9-Jun-15 |
| 280300 | Taus, Alvaro | CEIC Parc de Salut Mar | 9-Jun-15 |
| 280301 | Terrasa Pons, Josefa | CEIC Islas Baleares (CEIC-IB) | 9-Jun-15 |
| 280302 | Vazquez Estevez, Sergio | CEIC de Galicia (CAEI) | 9-Jun-15 |
| 280303 | Viñolas, Nuria | CEIC Hospital Clinic de Barcelona | 9-Jun-15 |
| 280506 | Ochsenbein, Adrian | Kantonale Ethikkommission Bern (KEK) | 3-Mar-16 |
| 280507 | Schütte, Wolfgang | Ethikkommission an der Universität Regensburg; Landshuter Straße 4 Raum 134, 1.OG. Regensburg 93047 Germany | 3-Nov-15 |
| 280508 | Stauder, Heribert | Ethikkommission an der Universität Regensburg; Landshuter Straße 4 Raum 134, 1.OG. Regensburg 93047 Germany | 3-Nov-15 |
| 280509 | Weißinger, Florian | Ethikkommission an der Universität Regensburg; Landshuter Straße 4 Raum 134, 1.OG. Regensburg 93047 Germany | 3-Nov-15 |
| 280510 | Engel-Riedel, Walburga | Ethikkommission an der Universität Regensburg; Landshuter Straße 4 Raum 134, 1.OG. Regensburg 93047 Germany | 3-Nov-15 |
| 280913 | Beatty, Patrick | Copernicus Group Independent Review Board; 1 Triangle Drive Suite 100 PO Box 110605 Research Triangle Park North Carolina 27709 United States | 17-Mar-15 |
| 280915 | Goueli, Basem | Saint Luke's Hospital Institutional Review Board; 915 East | 1-Sep-15 |

| Site # | Investigator | IRB/EC Name and Address | Approval Date |
|---|---|---|---|
| | | First Street Duluth Minnesota 55805 United States | |
| 280916 | Koh, Han | Kaiser Permanente Southern California Institutional Review Board; 393 East Walnut Street, 2nd Floor Pasadena California 91188 United States | 19-May-15 |
| 280917 | Schnell, Frederick | Copernicus Group Independent Review Board; 1 Triangle Drive Suite 100 PO Box 110605 Research Triangle Park North Carolina 27709 United States | 13-Jul-15 |
| 280919 | Nott, Louise | Tasmania Health and Medical Human Research Ethics Committee | 2-Jun-15 |
| 280920 | Yu, Chong-Jen | Research Ethics Committee of National Taiwan University Hospital | 16-Jun-15 |
| 280921 | Liu, Chien-Ying | Institutional Review Board of Chung Shan Medical University Hospital | 11-Jun-15 |
| 280922 | Greil, Richard | Ethikkommission für das Bundesland Salzburg; Sebastian-Stief-Gasse 2 Salzburg Salzburg 5010 Austria | 16-Oct-15 |
| 280925 | Reck, Martin | Ethikkommission an der Universität Regensburg; Landshuter Straße 4 Raum 134, 1.OG. Regensburg 93047 Germany | 3-Nov-15 |
| 280927 | Bourhaba, Maryam | CHU de Liège - Comité d'Ethique; Domaine Universitaire du Sart Tilman B35 Liege 4000 Belgium | 29-May-15 |
| 280928 | Goeminne, Jean-Charles | CHU de Liège - Comité d'Ethique; Domaine Universitaire du Sart Tilman B35 Liege 4000 Belgium | 29-May-15 |
| 280929 | Purkalne, Gunta | The Ethics Committee for Clinical Trials of Medicinal Products | 24-Apr-15 |
| 280930 | Lohri, Andreas | Kantonale Ethikkommission Bern (KEK) | 8-Mar-16 |
| 280932 | Hashemi, Sayed | Medical Research Ethics Committees United | 29-Oct-15 |
| 280983 | Masood, Nehal | Copernicus Group Independent Review Board; 1 Triangle Drive Suite 100 PO Box 110605 Research Triangle Park North Carolina 27709 United States | 4-Jun-15 |
| 280985 | Teixeira, Encarnação | Comissão de Ética para a Investigação Clínica - CEIC | 10-Jul-15 |
| 280986 | Adamchuk, Hryhoriy | CEQ of MI Kryvyi Rih Oncology Dispensary of Dnipropetrovsk Regional Council | 27-Feb-15 |
| 280987 | Andrusenko, Orest | CEQ of Treatment and Prevention Institution Volyn Regional Oncology Dispensary | 15-May-15 |
| 280988 | Bondarenko, Igor | CEQ of MI Dnipropetrovsk City Multifield Clinical Hospital #4 of Dnipropetrovsk Regional Council | 19-Mar-15 |
| 280989 | Chornobai, Anatolii | CEQ of Poltava Regional Clinical Oncology Dispensary of Poltava Regional Council | 26-Feb-15 |
| 280990 | Hotko, Yevhen | Commission on Ethics Questions of Uzhgorod Central City Clinical Hospital | 3-Mar-15 |
| 280991 | Ivashchuk, Oleksandr | Commission of Ethics Questions on the basis of the Chernivtsi Regional Clinical Oncology Dispensary | 8-Jul-15 |
| 280992 | Kolesnik, Oleksii | CEQ of MI of Zaporizhzhia Regional Council Zaporizhzhia Regional Clinical Oncology Dispensary | 11-Jun-15 |
| 280994 | Rusyn, Andriy | CEQ of Transcarpathian Regional Clinical Oncology Dispensary | 10-Mar-15 |
| 280995 | Vasylyev, Leonid | CEQ of SI Institute of Medical Radiology n.a. S.P. Hryhoriev of NAMS of Ukraine | 17-Mar-15 |
| 280996 | Shapovalov, Dmytro | CEQ of Municipal Noncommercial Institution Regional Center of Oncology | 20-Mar-15 |
| 280998 | Vynnychenko, Ihor | CEQ of Regional Municipal Institution Sumy Regional Clinical Oncology Dispensary | 2-Mar-15 |
| 280999 | Borra, Gloria | Comitato Etico Azienda Ospedaliera Universitaria Maggiore della Carità | 27-Jul-15 |
| 281001 | Barone, Carlo | Comitato Etico Dell Universita Cattolica del Sacro Cuore Policlinico Universitario Agostino Gemelli | 24-Sep-15 |
| 281002 | Girotto, Gustavo | Comitê de Ética em Pesquisa em Seres Humanos da Faculdade de Medicina de São José do Rio Preto | 11-May-15 |
| 281003 | Aragao, Bruno | Comitê de Ética em Pesquisa em Seres Humanos do Hospital Socor | 9-Mar-16 |
| 281005 | Faccio, Adilson | Comitê de Ética em Pesquisa em Seres Humanos da Universidade de Ribeirão Preto (UNAERP) | 13-Apr-16 |
| 281009 | Campos, Clodoaldo | Comitê de Ética em Pesquisa em Seres Humanos da | 8-May-16 |

| Site # | Investigator | IRB/EC Name and Address | Approval Date |
|---|---|---|---|
| | | Irmandade da Santa Casa de Londrina | |
| 281010 | Matias, Danielli | Comitê de Ética em Pesquisa em Seres Humanos da Liga Norte Riograndense Contra o Câncer | 2-Feb-16 |
| 281011 | da Silva, Carlos | Comitê de Ética em Pesquisa Fundação Pio XII Hospital de Câncer de Barretos | 18-Feb-16 |
| 281013 | Lopez, Yamil | Comité de Ética en Investigación de la Facultad de Medicina y Hospital Universitario | 12-Aug-15 |
| 281857 | Faller, Bryan | Missouri Baptist Medical Center Institutional Review Board; 3015 North Ballas Road St. Louis Missouri 63131 United States | 13-Jul-15 |
| 281858 | Paschold, John | US Oncology Inc. Institutional Review Board; 10101 Woodloch Forest The Woodlands Texas 77380 United States | 18-Jun-15 |
| 281860 | Ou, Sai-Hong Ignatius | University of California Irvine Institutional Review Board; 5171 Calfornia Avenue, Office Of Research Suite 150 Irvine California 92697 United States | 22-Jul-15 |
| 281904 | Carr, Laurie | Western Institutional Review Board; 1019 39th Avenue Southeast Suite 120 Puyallup Washington 98374 United States | 16-Oct-15 |
| 281905 | Daniels, Gregory | University of California, San Diego Human Research Protections Program; 3350 La Jolla Village Drive San Diego California 92161 United States | 18-Feb-16 |
| 281906 | Eskander, Elhamy | Frederick Memorial Hospital Institutional Review Board; 400 West 7th Street Frederick Maryland 21701 United States | 27-Jun-16 |
| 281907 | Kotiah, Sandy | Mercy Medical Center IRB; 345 St. Paul Place Bunting Center, 7th Floor Baltimore Maryland 21202 United states | 20-Apr-15 |
| 281908 | Kuzma, Charles | Copernicus Group Independent Review Board; 1 Triangle Drive Suite 100 PO Box 110605 Research Triangle Park North Carolina 27709 United States | 19-May-15 |
| 281909 | Hoffman, Philip | University of Chicago Hospitals Institutional Review Board; 5751 South Woodlawn Avenue McGiffert Hall Chicago Illinois 60637 United States | 7-Aug-15 |
| 281910 | Rodriguez, Estelamari | Mount Sinai Medical Center IRB; 4300 Alton Road Miami Beach Florida 33140 United States | 19-Oct-15 |
| 281944 | Pastor, Andrea | Comité Provincial de Bioética - Ministerio de Salud de la Provincia de Santa Fé | 28-Oct-15 |
| 281945 | Streich, Guillermo | Comité Independiente de Etica en investigación clínica "Dr. Carlos A. Barclay | 30-Jun-15 |
| 281946 | Aerts, Joachim | Medical Research Ethics Committees United | 29-Oct-15 |
| 281947 | Schramel, Franz | Medical Research Ethics Committees United | 29-Oct-15 |
| 281948 | Snijders, Dominic | Medical Research Ethics Committees United | 29-Oct-15 |
| 281950 | Aerts, Joachim | Medical Research Ethics Committees United | 29-Oct-15 |
| 281954 | Dickgreber, Nicolas | Ethikkommission an der Universität Regensburg; Landshuter Straße 4 Raum 134, 1.OG. Regensburg 93047 Germany | 3-Nov-15 |
| 281957 | Kollmeier, Jens | Ethikkommission an der Universität Regensburg; Landshuter Straße 4 Raum 134, 1.OG. Regensburg 93047 Germany | 3-Nov-15 |
| 281964 | Gomez-Villanueva, Angel | Comite de Etica Investigacion de la Clinica Bajío | 5-Jun-15 |
| 281966 | Dominguez Andrade, Adriana | Comite de Etica en Investigacion de Mexico Centre for Clinical Research SA de CV | 15-May-15 |
| 281966 | Dominguez Andrade, Adriana | Comite de Etica en Investigacion de Mexico Centre for Clinical Research SA de CV | 15-May-15 |
| 281983 | Costamilan, Rita de Cassia | Comitê de Ética em Pesquisa da Universidade de Caxias do Sul | 29-Mar-16 |
| 282032 | Schwartsmann, Gilberto | Comitê de Ética em Pesquisa do Hospital de Clínicas de Porto Alegre | 6-Apr-16 |
| 282033 | Tadokoro, Hakaru | Comite de Etica em Pesquisa da Universidade Federal de Sao Paulo - Hospital Sao Paulo | 17-Mar-16 |
| 282034 | Bruno, Luiz | Comitê de Ética em Pesquisa - Hospital Mãe de Deus | 10-Mar-16 |
| 282035 | Abbade Dettino, Aldo | Comitê de Ética em Pesquisa da Fundação Antônio Prudente – Hospital do Câncer A. C. Camargo | 6-May-16 |

| Site # | Investigator | IRB/EC Name and Address | Approval Date |
|---|---|---|---|
| 282036 | Dimitrov, Borislav | Ethics Committee for Multi-Centre Trials; 5 Sveta Nedelya Square Sofia Sofia-Grad 1000 Bulgaria | 3-Jun-15 |
| 282037 | Koynov, Krassimir | Ethics Committee for Multi-Centre Trials; 5 Sveta Nedelya Square Sofia Sofia-Grad 1000 Bulgaria | 3-Jun-15 |
| 282038 | Mihaylova, Zhasmina | Ethics Committee for Multi-Centre Trials; 5 Sveta Nedelya Square Sofia Sofia-Grad 1000 Bulgaria | 20-Jan-16 |
| 282039 | Ilieva, Rumyana | Ethics Committee for Multi-Centre Trials; 5 Sveta Nedelya Square Sofia Sofia-Grad 1000 Bulgaria | 3-Jun-15 |
| 282043 | Galiulin, Rinat | Ethics Committee at Clinical Oncology Dispensary | 22-Jun-15 |
| 282044 | Karaseva, Nina | Ethics Committee at City Clinical oncologic dispensary | 7-Jul-15 |
| 282047 | Gorbunova, Vera | Ethics Committee at Russian Oncology Research Center n.a. N.N.Blokhin | 30-Jun-15 |
| 282048 | Stroyakovskii, Daniil | Ethics Committee at Moscow City Oncology Hospital #62 of Moscow Healthcare Department | 26-Jul-15 |
| 282049 | Kovalenko, Nadezhda | Ethics Committee at Volzhskiy regional clinical oncology dispensary #3 | 27-Jun-16 |
| 282051 | Berard, Henri | CPP Sud-Méditerranée 2; 270 Boulevard Sainte Marguerite Hôpital Sainte Margeurite Pavillon 9 Cedex 9 Marseille Bouches-du-Rhône 13274 France | 11-Jun-15 |
| 282053 | Coudert, Bruno | CPP Sud-Méditerranée 2; 270 Boulevard Sainte Marguerite Hôpital Sainte Margeurite Pavillon 9 Cedex 9 Marseille Bouches-du-Rhône 13274 France | 5-Jun-15 |
| 282054 | Denis, Fabrice | CPP Sud-Méditerranée 2; 270 Boulevard Sainte Marguerite Hôpital Sainte Margeurite Pavillon 9 Cedex 9 Marseille Bouches-du-Rhône 13274 France | 5-Jun-15 |
| 282056 | Fabre, Elizabeth | CPP Sud-Méditerranée 2; 270 Boulevard Sainte Marguerite Hôpital Sainte Margeurite Pavillon 9 Cedex 9 Marseille Bouches-du-Rhône 13274 France | 5-Jun-15 |
| 282163 | Barlesi, Fabrice | CPP Sud-Méditerranée 2; 270 Boulevard Sainte Marguerite Hôpital Sainte Margeurite Pavillon 9 Cedex 9 Marseille Bouches-du-Rhône 13274 France | 5-Jun-15 |
| 282164 | Dayen, Charles | CPP Sud-Méditerranée 2; 270 Boulevard Sainte Marguerite Hôpital Sainte Margeurite Pavillon 9 Cedex 9 Marseille Bouches-du-Rhône 13274 France | 5-Jun-15 |
| 282165 | Foa, Cyril | CPP Sud-Méditerranée 2; 270 Boulevard Sainte Marguerite Hôpital Sainte Margeurite Pavillon 9 Cedex 9 Marseille Bouches-du-Rhône 13274 France | 5-Jun-15 |
| 282166 | Gazaille, Virgile | CPP Sud-Méditerranée 2; 270 Boulevard Sainte Marguerite Hôpital Sainte Margeurite Pavillon 9 Cedex 9 Marseille Bouches-du-Rhône 13274 France | 5-Jun-15 |
| 282167 | Veillon, Remi | CPP Sud-Méditerranée 2; 270 Boulevard Sainte Marguerite Hôpital Sainte Margeurite Pavillon 9 Cedex 9 Marseille Bouches-du-Rhône 13274 France | 5-Jun-15 |
| 282168 | Morere, Jean François | CPP Sud-Méditerranée 2; 270 Boulevard Sainte Marguerite Hôpital Sainte Margeurite Pavillon 9 Cedex 9 Marseille Bouches-du-Rhône 13274 France | 5-Jun-15 |
| 282169 | Audigier-Valette, Clarisse | CPP Sud-Méditerranée 2; 270 Boulevard Sainte Marguerite Hôpital Sainte Margeurite Pavillon 9 Cedex 9 Marseille Bouches-du-Rhône 13274 France | 5-Jun-15 |
| 282213 | Kryzhanivska, Anna | CEQ of Ivano-Frankivsk Regional Oncology Dispensary | 12-May-16 |
| 282442 | Rodriguez-Abreu, Delvys | CEIC Hospital Universitario Insular Materno-Infantil de Las Palmas | 9-Jun-15 |
| 282444 | Felip Font, Enriqueta | CEIC Hospital Universitari Vall d'Hebron | 9-Jun-15 |
| 282445 | Garrido Lopez, Pilar | CEIC Hospital Universitario Ramon y Cajal | 9-Jun-15 |
| 282447 | Ponce Aix, Santiago | CEIC Hospital Universitario 12 de Octubre | 9-Jun-15 |
| 282448 | Beniak, Juraj | Eticka komisia Presovskeho samospravneho kraja | 29-Jun-15 |
| 282449 | Kasan, Peter | Eticka komisia Univerzitna nemocnica Bratislava | 29-Jun-15 |
| 282452 | Cicenas, Saulius | Lithuanian Bioethics Committee | 20-May-15 |
| 282454 | Filipauskiene, Judita | Lithuanian Bioethics Committee | 20-May-15 |

| Site # | Investigator | IRB/EC Name and Address | Approval Date |
|---|---|---|---|
| 282455 | Nadal, Ernest | CEIC Hospital Universitari de Bellvitge | 9-Jun-15 |
| 282880 | Godal, Robert | Eticka komisia pri Narodnom onkologickom ustave | 29-Jun-15 |
| 282882 | Moron Escobar, Hernan | Asociacion Benefica Prisma | 28-May-15 |
| 282884 | Gurubhagavatula, Sarada | Copernicus Group Independent Review Board; 1 Triangle Drive Suite 100 PO Box 110605 Research Triangle Park North Carolina 27709 United States | 20-May-15 |
| 282885 | Nissenblatt, Michael | Copernicus Group Independent Review Board; 1 Triangle Drive Suite 100 PO Box 110605 Research Triangle Park North Carolina 27709 United States | 19-May-15 |
| 282886 | Herman, James | WIRB Copernicus Group; 1 Triangle Drive Suite 100 PO Box 110605 Research Triangle Park North Carolina 27709 United States | 29-Jan-16 |
| 282887 | Kono, Scott | Kaiser Permanente of Colorado Institutional Review Board; 10065 East Harvard Avenue Suite 300 Denver Colorado 80231 United States | 27-Jan-16 |
| 282970 | Zhiltsova, Elena | Ethics Committee at Russian Medical Military Academy n.a. S.M.Kirov | 30-Jun-15 |
| 283383 | Cisneros Tipismana, Rocio | Comite de Etica en Investigacion del Instituto Regional de Enfermedades Neoplasicas | 13-Nov-15 |
| 284076 | Su, Wu-Chou | National Cheng Kung University Hospital Human Experiment and Ethic Committee | 12-Jun-15 |
| 284111 | VanderWalde, Ari | Western Institutional Review Board (WIRB); 1019 39th Avenue Southeast Suite 120 Puyallup Washington 98374 United States | 16-Nov-15 |
| 284113 | Bernicker, Eric | Houston Methodist Research Institute IRB; 6670 Bertner Suite 6-351 Houston Texas 77030 United States | 15-Dec-15 |
| 284120 | Socoteanu, Matei | US Oncology Inc. Institutional Review Board; 10101 Woodloch Forest The Woodlands Texas 77380 United States | 18-jun-15 |
| 284122 | Fiorillo, Joseph | US Oncology Inc. Institutional Review Board; 10101 Woodloch Forest The Woodlands Texas 77380 United States | 18-jun-15 |
| 284123 | Kozloff, Mark | Ingalls Memorial Hospital IRB; 1 Ingalls Drive Ingalls Memorial Hospital Harvey Illinois 60426 United States | 22-Jul-15 |
| 284126 | Grossi, Francesco | Comitato Etico San Martino IST 2 | 11-Mar-16 |
| 284128 | Dingemans, Anne-Marie | Medical Research Ethics Committees United | 29-Oct-15 |
| 284130 | Mazieres, Julien | CPP Sud-Méditerranée 2; 270 Boulevard Sainte Marguerite Hôpital Sainte Margeurite Pavillon 9 Cedex 9 Marseille Bouches-du-Rhône 13274 France | 5-Jun-15 |
| 284672 | Braiteh, Fadi | Copernicus Group Independent Review Board; 1 Triangle Drive Suite 100 PO Box 110605 Research Triangle Park North Carolina 27709 United States | 4-Aug-15 |
| 284673 | Narang, Mohit | US Oncology Inc. Institutional Review Board; 10101 Woodloch Forest The Woodlands Texas 77380 United States | 18-jun-15 |
| 284674 | Spira, Alexander | US Oncology Inc. Institutional Review Board; 10101 Woodloch Forest The Woodlands Texas 77380 United States | 18-Jun-15 |
| 284675 | Batus, Marta | Rush University Medical Center Institutional Review Board; 1653 West Congress Parkway Chicago Illinois 60612 United States | 15-Jan-16 |
| 285270 | Tummala, Mohan | Mercy Health Springfield Communities Institutional Review Board; 1235 East Cherokee Street Springfield Missouri 65804 United States | 21-Aug-16 |
| 285271 | Jhangiani, Haresh | Copernicus Group Independent Review Board; 1 Triangle Drive Suite 100 PO Box 110605 Research Triangle Park North Carolina 27709 United States | 16-Jul-15 |
| 285273 | Menefee, Michael | Mayo Clinic Institutional Review Board; 200 First Street Southwest Rochester Minnesota 55905 United States | 10-Aug-16 |
| 285274 | Rafiyath, Shamudheen | Salus IRB; 2111 West Braker Lane Suite 400 Austin Texas 78758 United States | 11-Dec-15 |
| 286362 | Hsieh, Ruey-Kuen | Mackay Memorial Hospital Institutional Review Board | 17-Aug-15 |

| Site # | Investigator | IRB/EC Name and Address | Approval Date |
|---|---|---|---|
| 288218 | Shamrai, Volodymyr | Commission on Ethics Questions of Vinnytsya Regional Clinical Oncology Dispensary | 22-Jul-15 |
| 288407 | Orellana Ulunque, Eric | Comite Etico Cientifico Clinica Santa Maria | 19-Oct-15 |
| 288408 | Cheng, Haiying | BRANY IRB; 225 Community Drive Suite 100 Great Neck New York 11021 United States | 1-Dec-15 |
| 288409 | Eisenberg, Peter | Copernicus Group Independent Review Board; 1 Triangle Drive Suite 100 PO Box 110605 Research Triangle Park North Carolina 27709 United States | 14-Aug-15 |
| 288410 | Sabbath, Kert | Yale University Human Research Protection Program; 55 College Street New Haven Connecticut 6510 United States | 2-Mar-16 |
| 288811 | Becker, Kevin | Maimonides Med Ctr Institutional Review Board; 4802 10th Avenue Brooklyn New York 11219 United States | 1-Sep-15 |
| 288812 | Kosty, Michael | Scripps Health Institutional Review Board; 11025 North Torrey Pines Road Suite 200 La Jolla California 92037 United States | 7-Mar-16 |
| 288813 | Fabregas, Jesus | Copernicus Group Independent Review Board; 1 Triangle Drive Suite 100 PO Box 110605 Research Triangle Park North Carolina 27709 United States | 17-Nov-15 |
| 288814 | Karnad, Anand | University of Texas Health Science Center San Antonio Institutional Review Board; 7703 Floyd Curl Drive Greyhound North Campus Research Administration Room 2.104 San Antonio Texas 78229 United States | 20-Oct-15 |
| 288815 | Joshi, Abhishek | Melbourne Health Human Research Ethics Committee | 18-Nov-15 |
| 291888 | Zylla, Dylan | Park Nicollet Institute Institutional Review Board; 3800 Park Nicollet Boulevard Minneapolis Minnesota 55416 United States | 18-mar-16 |
| 292837 | Chovanec, Jozef | Eticka komisia NsP Sv. Jakuba, n.o., Bardejov | 2-Jun-16 |
| 292838 | Steppert, Claus | Ethikkommission der Bayerischen Landesärztekammer | 15-Jun-16 |
| 292994 | Ogorodnikova, Nina | CEQ of Kyiv City Clinical Oncological Center | 18-Jul-16 |
| 293115 | Hsia, Te-Chun | The Institutional Review Board of China Medical University Hospital | 3-Jun-16 |
| 293119 | Penkov, Konstantin | Ethics Committee at Private Medical Institution "Evromedservis" | 27-May-16 |
| 293120 | Areses Manrique, Maria del Carmen | CEIC de la Corporacion Sanitaria del Parc Tauli | 26-Apr-16 |
| 293124 | Hackanson, Björn | Ethikkommission der Bayerischen Landesärztekammer | 15-Jun-16 |
| 293125 | Gironés, Regina | CEIC de la Corporacion Sanitaria del Parc Tauli | 26-Apr-16 |
| 293130 | Gautschi, Oliver | Kantonale Ethikkommission Bern (KEK) | 2-Aug-16 |
| 293132 | Lammers, Ernst | Medical Research Ethics Committees United | 13-Jun-16 |
| 293294 | Le Moulec, Sylvestre | CPP Sud-Méditerranée 2; 270 Boulevard Sainte Marguerite Hôpital Sainte Margeurite Pavillon 9 Cedex 9 Marseille Bouches-du-Rhône 13274 France | 13-May-16 |
| 293295 | Barlo, Nicole | Medical Research Ethics Committees United | 13-Jun-16 |
| 293296 | Keizer - van 't Westeinde, Susan | Medical Research Ethics Committees United | 13-Jun-16 |
| 293301 | van Haarst, Jan Maarten | Medical Research Ethics Committees United | 13-Jun-16 |
| 293338 | Pazzola, Antonio | Comitato di Bioetica dell'AUSL 1 di Sassari | 20-Jun-16 |
| 293747 | Niederman, Thomas | Copernicus Group Independent Review Board; 1 Triangle Drive Suite 100 PO Box 110605 Research Triangle Park North Carolina 27709 United States | 3-May-16 |
| 293748 | Raez, Luis | Western Institutional Review Board; 1019 39th Avenue Southeast Suite 120 Puyallup Washington 98374 United States | 30-Sep-16 |
| 293749 | Porubska, Miriam | Eticka komisia Onkologicky ustav sv. Alzbety | 2-Jun-16 |
| 293750 | Vasiliev, Aleksandr | Ethics Committee at Railway Clinical Hospital JSC RZhD | 27-May-16 |
| 293751 | Koleva, Marchela | Ethics Committee for Multi-Centre Trials; 5 Sveta Nedelya Square Sofia Sofia-Grad 1000 Bulgaria | 29-Aug-2016 |

| Site # | Investigator | IRB/EC Name and Address | Approval Date |
|---|---|---|---|
| 293752 | van Lindert, Anne | Medical Research Ethics Committees United | 13-Jun-16 |
| 293753 | Mansour, Khaled | Medical Research Ethics Committees United | 13-Jun-16 |
| 294212 | Rosales, Joseph | Western Institutional Review Board; 1019 39th Avenue Southeast Suite 120 Puyallup Washington 98374 United States | 20-Jul-16 |
| 294298 | Spadafora, Silvana | Sault Area Hospital Research Ethics Board; 750 Great Northern Road Sault Ste. Marie Ontario P6B 0A8 Canada | 23-Jun-16 |
| 294828 | Naidu, Sashi | Copernicus Group Independent Review Board; 1 Triangle Drive Suite 100 PO Box 110605 Research Triangle Park North Carolina 27709 United States | 8-Jul-16 |
| 294829 | Lee, Arielle | Copernicus Group Independent Review Board; 1 Triangle Drive Suite 100 PO Box 110605 Research Triangle Park North Carolina 27709 United States | 20-Jun-16 |
| 294830 | Matthews-Smith, Velmalia | Copernicus Group Independent Review Board; 1 Triangle Drive Suite 100 PO Box 110605 Research Triangle Park North Carolina 27709 United States | 1-Jul-16 |
| 294833 | Saturnino Duarte de Brito, Ulisses | Comissão de Ética para a Investigação Clínica - CEIC | 27-Sep-16 |
| 295101 | Keogh, George | Copernicus Group Independent Review Board; 1 Triangle Drive Suite 100 PO Box 110605 Research Triangle Park North Carolina 27709 United States | 6-Sep-16 |
| 285227 | Kishi, Kazuma | Toranomon Hospital and Toranomon Hospital Kajigaya IRB, 2-2-2 Toranomon, Minato-ku, 105-8470, Tokyo, JAPAN | 14-Jul-2015 |
| 285229 | Otani, Sakiko | Kitasato University Sagamihara IRB, 1-15-1 Kitasato, Minami-ku, Sagamihara-shi, 252-0375, Kanagawa, JAPAN | 15-Jul-2015 |
| 285230 | Tanaka, Hiroshi | Niigata Cancer Center Hospital IRB, 2-15-3 Kawagishi-cho, Chuo-ku, Niigata-shi, 951-8566, Niigata, JAPAN | 13-Jul-2015 |
| 285231 | Kim, Young Hak | Kyoto University Hospital IRB, 54 Kawahara-cho Shogoin Sakyo-ku, Kyoto-shi, 606-8507, Kyoto, JAPAN | 15-Jul-2015 |
| 285232 | Kawaguchi, Tomoya | Osaka City University Hospital IRB, 1-5-7,Asahimachi, Abeno-ku, Osaka-shi, 545-8586, Osaka, JAPAN | 22-Jul-2015 |
| 285233 | Yokota, Soichiro | National Hosptial Organization Toneyama National Hospital IRB, 5-1-1 Toneyama, Toyonaka-shi, 560-8552, Osaka, JAPAN | 31-Jul-2015 |
| 285234 | Yamamoto, Nobuyuki | Wakayama Medical University IRB, 811-1 Kimiidera, Wakayama-shi, 641-8509, Wakayama, JAPAN | 21-Jul-2015 |
| 285235 | Azuma, Koichi | Kurume University IRB, 67 Asahimachi, Kurume-shi, Fukuoka, 830-0011, JAPAN | 21-Jul-2015 |
| 285236 | Seto, Takashi | National Hospital Organization Kyushu Cancer Center; IRB, 3-1-1 Notame, Minami-ku, Fukuoka-shi, 811-1395, Fukuoka, JAPAN | 1-Jul-2015 |
| 285300 | Ikeda, Satoshi | Kanagawa Cardiovascular and Respiratory Center IRB, 6-16-1 Tomiokahigashi, Kanazawa-ku, Yokohama-shi, 236-0051, Kanagawa, JAPAN | 14-Jul-2015 |
| 285301 | Ichiki, Masao | National Hospital Organization Kyushu Medical Center IRB, 1-8-1 Jigyohama, Chuo-Ku, Fukuoka-shi, 810-8563, Fukuoka, JAPAN | 22-Jul-2015 |
| 285302 | Nogami, Naoyuki | National Hospital Organization Shikoku Cancer Center IRB, 160 Minamiumemotomachi-Kou, Matsuyama-shi, 791-0280, Ehime, JAPAN | 23-Jul-2015 |
| 286439 | Fukuhara, Tatsuro | Miyagi Cancer Center IRB, 47-1 Nodayama, Medeshima-Shiote, Natori-shi, 981-1293, Miyagi, JAPAN | 15-Sep-2015 |
| 286754 | Yokoyama, Takuma | Kyorin University Hospital IRB, 6-20-2 Shinkawa, Mitaka-shi, 181-8611, Tokyo, JAPAN | 12-Aug-2015 |
| 286755 | Takeda, Yuichiro | Center Hospital of the National Center for Global Health and Medicine IRB, 1-21-1 Toyama, Shinjuku-ku, 162-8655, Tokyo, Japan | 16-Jul-2015 |

### IMpower130

| Country | EC/IRB | Address |
|---|---|---|
| Belgium | Cliniques Universitaires Saint-Luc - Comité | Promenade de l'Alma 51 bte B1.43.03 Bruxelles 1200 Belgium |
| Canada | Comite d'ethique de la recherche de l' Hopital Maisonneuve-Rosemont | 10 Rue De L'Espinay Quebec Quebec G1L 3L5 Canada |
| Canada | McGill University Health Center Montreal Hospital | 1032 W. Sheridan Road Research Services Granada Center, Suite 400 Chicago Illinois 60660 United States |
| Canada | Royal Victoria Regional Health Centre Research Ethics Board | Royal Perth Hospital, Wellington Street Perth Washington 6001 Australia |
| Canada | UBC BCCA Research Ethics Board | 750 West Broadway Fairmont Medical Building Suite 902 Vancouver British Columbia V5Z 1H5 Canada |
| Canada | William Osler Health system Research Ethics Board | 2100 Bovaird Drive East Brampton Ontario L6W 3J7 Canada |
| France | CPP Est III | 1020 Almira Street 3rd Floor, Room 321 Saginaw Michigan 48602 United States |
| Germany | Ethikkommission der Landesärztekammer Baden-Württemberg | Bachstrasse 18 Gebaeude 1 Jena 7740 Germany |
| Germany | Ethik-Kommission der Sächsischen Landesärztekammer | Gartenstrasse 210 - 214 Münster Nordrhein-Westfalen 48147 Germany |
| Hong Kong | Joint Chinese University of Hong Kong - New Territories East Cluster Clinical Research Ethics | 30 - 32 Ngan Shing Street, Prince of Wales Hospital 8th Floor, Lui Che Woo Clinical Science Building, Hong Kong Hong Kong |
| Israel | Assaf Harofe Medical Center EC | Beer Yaakov 70300 Zerifin 70300 Israel |
| Israel | Galilee Medical Center EC | Western Galilee Hospital POB 21 Nahariya 22100 Israel |
| Israel | Hadassah University Hospital Local EC | Wilgenstraat 2 Campus Wilgenstraat Roeselare West-Vlaanderen 8800 Belgium |
| Israel | Kaplan Medical Center Local EC | Kaplan Medical Center P.o Box 1 Rehovot 76100 Israel |
| Israel | Meir EC | National Institute Of Health, C/o Institute For Health Management, Kuala Lumpur Bangsar WilayahPersekutuan KualaLumpur 59000 Malaysia |
| Israel | Rabin Medical Center Ethics Committee | 1501 Fourth Avenue Suite 800 Seattle Washington 98101 United States |
| Israel | Rabin Medical Center Local EC | 39 Jabutinsky street Petah Tikva 49100 Israel |
| Israel | Rambam Medical Center Ethics Committee | 8 Haaliya Hashniya Street Bat Galim Haifa 31096 Israel |
| Israel | Soroka University Medical Center Local EC | Post Office Box 151 Beer Sheva 84101 Israel |
| Israel | Tel Aviv Sourasky EC | 6 Weitzman Street Tel Aviv Tel-Aviv 64239 Israel |
| Israel | The Chaim Sheba Medical Center EC | Chaim Sheba Medical Center Tel Hashomer Ramat Gan 52621 Israel |
| Italy | Comitato Etico Campania Nord | Via Giovanbattista Pergolesi 33 Monza Lombardia 20900 Italy |
| Italy | Comitato Etico delle Aziende Sanitarie dell'Umbria | Largo Francesco Vito, 1 Roma Lazio 168 Italy |

| | | |
|---|---|---|
| **Italy** | **Comitato Etico delle Province di Chieti e Pescara** | Largo Francesco Vito, 1 Roma Lazio 168 Italy |
| **Italy** | **Comitato Etico IRCCS**<br>**Istituto Nazionale per lo Studio e la Cura dei Tumori Fondazione G Pascale** | Via Dell'eremo 9/11 Lecco Lombardia 23900 Italy |
| **Italy** | **Comitato Etico Regionale delle Marche** | Via Olgettina, 60 Milano Lombardia 20132 Italy |
| **Italy** | **Comitato Etico Seconda Università degli Studi di Napoli Az. Osp. Univ. S.U.N. - A.O.R.N. "Ospedali** | Via Olgettina, 60 Milano Lombardia 20132 Italy |
| **Spain** | **CEIC Consorcio Hospital General Universitario de Valencia** | Avenida Tres Cruces, 2 Consorcio Hospital General Universitario de Valencia Pabellon B-3 - 4ª planta Valencia Valencia 46014 Spain |
| **Spain** | **CEIC de Aragon (CEICA)** | Calle Micer Masco, 31 Valencia Valencia 46010 Spain |
| **Spain** | **CEIC de Galicia (CAEI)** | Calle Micer Masco, 31 Valencia Valencia 46010 Spain |
| **Spain** | **CEIC Hospital de la Santa Creu i Sant Pau** | Avenida Vicente Blasco Ibánez, 17 Valencia Valencia 46010 Spain |
| **Spain** | **CEIC Hospital Santa Creu i Sant Pau** | Avenida Sant Antoni Maria Claret, 167 Servicio de Famacología Clínica Pabellón HC Barcelona Barcelona 8025 Spain |
| **Spain** | **CEIC Hospital Universitario de Canarias** | Avda. de Córdoba s/n Instituto de Investigación Hospital 12 de Octubre (i+12) Bloque D - Planta 6ª Area de Gestión de Proyectos - Unidad Administrativa CEIC Madrid 28041 Spain |
| **United States** | **Appalachian Regional Healthcare IRB** | 58 Canal Circular Road Apollo Gleneagles Hospitals Kolkata Kolkata West Bengal 700054 India |
| **United States** | **Banner MD Anderson Cancer Center IRB** | 2940 E. Banner Gateway Dr. Suite 375 Gilbert California 85234 United States |
| **United States** | **Biomedical Research Alliance of New York LLC Institutional Review Board** | University Of North Carolina, Cb # 7097 Medical School Building 52 Chapel Hill North Carolina 27599 United States |
| **United States** | **Birmingham Veterans Administration Medical Center IRB** | 207 Olds Hall East Lansing Michigan 48824 United States |
| **United States** | **Copernicus Group Independent Review Board** | 5000 CentreGreen Way Suite 200 Cary North Carolina 27513 United States |
| **United States** | **Copernicus IRB** | 1 Triangle Drive Suite 100 Po Box 110605 Research Triangle Park North Carolina 27709 United States |
| **United States** | **Duke University Health System Institutional Review Board** | Hock Plaza Durham North Carolina 27705 United States |
| **United States** | **Englewood Hospital and Medical Center** | 350 Engle Street Englewood New Jersey 7631 United States |
| **United States** | **Kaiser Permanente Northern California Institutional Review Board** | 1800 Harrison Street 16th Floor Oakland California 94162 United States |

| | | |
|---|---|---|
| **United States** | **Kaiser Permanente of Colorado Institutional Review Board** | 3800 North Interstate Avenue Portland Oregon 97227 United States |
| **United States** | **Lahey Clinic, Inc. Institutional Review Board** | 3-23-1, Shiobara, Minami-ku Fukuoka-shi 815-8588 Japan |
| **United States** | **Lancaster General Hospital IRB** | 3-23-1, Shiobara, Minami-ku Fukuoka-shi 815-8588 Japan |
| **United States** | **Loyola University Institutional Review Board** | 1032 W. Sheridan Road Research Services Granada Center, Suite 400 Chicago Illinois 60660 United States |
| **United States** | **Mayo Clinic Institutional Review Board** | 1032 W. Sheridan Road Research Services Granada Center, Suite 400 Chicago Illinois 60660 United States |
| **United States** | **MD Anderson Institutional Review Board** | 1400 Pressler Street Unit 1452 Houston Texas 77030-4009 United States |
| **United States** | **New England Institutional Review Board** | 85 Wells Avenue Suite 107 Newton Massachussets 2459 United States |
| **United States** | **North Mississippi Health Services** | 830 South Gloster Street Tupelo Mississippi 38801 United States |
| **United States** | **NYU School of Medicine Institutional Review Board** | 68, Hangeulbiseok-ro,Nowon-gu Seoul 1830 Korea, Republic of |
| **United States** | **Ochsner Clinic Foundation Institutional Review** | 68, Hangeulbiseok-ro,Nowon-gu Seoul 1830 Korea, Republic of |
| **United States** | **Pinnacle Health Hospitals Institutional Review** | 1968 Peachtree Road Northwest Atlanta Georgia 30309 United States |
| **United States** | **Rhode Island Hospital Institutional Review Board** | 1 Hoppin Street Office of Research Administration, Coral West Suite 1300 Committee on the Protection of Human Subjects Providence Rhode Island 2903 United States |
| **United States** | **Saint Barnabas Medical Center Institutional Review Board** | 1055 North Curtis Road Boise Idaho 83706 United States |
| **United States** | **Siouxland Institutional Review Board** | 230 Nebraska Street Sioux City Iowa 51101 United States |
| **United States** | **Springfield Committee for Research Involving Human Subjects (SCRIHS)** | 801 North Rutledge Street Springfield Illinois 62702 United States |
| **United States** | **The Christ Hospital IRB** | 3535 Market Street Suite 1200 Philadelphia Pennsylvania 19104 United States |
| **United States** | **University of Arkansas IRB** | Lembah Pantai Kuala Lumpur 59100 Malaysia |
| **United States** | **University of Chicago Hospitals Institutional Review Board** | 5751 South Woodlawn Avenue McGiffert Hall Chicago Illinois 60637 United States |
| **United States** | **University Of Iowa Human Subjects Office IRB** | 1 Illini Drive Peoria Illinois 61656 United States |
| **United States** | **University of Louisville IRB** | 501 East Broadway Medcenter One Suite 200 Louisville Kentucky 40202 United States |
| **United States** | **University Of Miami** | 55 Lake Ave., North Worcester Massachussets 1655 United States |
| **United States** | **University of Nevada Reno Research Integrity Office** | 987830 Nebraska Medical Center Omaha Nebraska 68198-7830 United States |
| **United States** | **W.G. 'Bill' Hefner VA Medical Center** | 1601 Brenner Avenue Salisbury North Carolina 28144 United States |

| | | |
|---|---|---|
| **United States** | **Walter Reed National Military Medical Center IRB** | 1601 Brenner Avenue Salisbury North Carolina 28144 United States |
| **United States** | **Western Institutional Review Board** | 1019 39th Avenue Southeast Suite 120 Puyallup Washington 98374 United States |
| **United States** | **Western Institutional Review Board (WIRB)** | 1019 39th Avenue Southeast Suite 120 Puyallup Washington 98374 United States |
| **United States** | **WIRB Copernicus Group** | 222 Station Plaza North Suite 521 Mineola New York 11501 United States |

### IMpower131

| Site # | Country | Investigator | EC/IRB Name and Address | IRB/EC Approval |
|---|---|---|---|---|
| 279803 | Argentina | Jarchum, Gustavo | Comité Institutcional de Etica de Investigación en Salud Sanatorio Allende, Hipolito Yrigoyen 384, X5000JHQ, Córdoba, Córdoba, Argentina | 11/12/15 |
| 279804 | Argentina | Magri, Ignacio | Comité Institucional de Ética de Investigación en Salud del Instituto Médico Río Cuarto, Hipólito Yrigoyen 1020, 5800, Rio Cuarto, Córdoba, Argentina | 10/6/15 |
| 282502 | Argentina | Pastor, Andrea | Comite De Etica Del Hospital Provincial Del Centenario, Urquiza 3101, S2002KDS, Rosario, Santa Fe, Argentina | 11/25/15 |
| 279805 | Argentina | Kotliar, Mauricio | Comité de Ética Independiente - Fundación Sanatorio, Francisco Acuña de Figueroa 1240, C1180AAX, Buenos Aires, Ciudad Autónoma de BuenosAires, Argentina | 4/28/15 |
| 279801 | Argentina | Cigno, Edgardo | Comite De Etica Del Sanatorio Britanico Sa, Paraguay 40, S2000CVB, Rosario, Santa Fe, Argentina | 1/26/16 |
| 279800 | Argentina | Martin, Claudio | Comité de Etica en Investigación del Instituto Alexander Fleming, Cramer 1180, C1426ANZ, Buenos Aires, Ciudad Autónoma de BuenosAires, Argentina | 9/16/15 |
| 279799 | Argentina | Lerzo, Guillermo | Comité Independiente de Etica en investigación clínica "Dr. Carlos A. Barclay, Larrea 1381, 3° A, 1117, Buenos Aires, Ciudad Autónoma de BuenosAires, Argentina | 2/12/15 |
| 279798 | Argentina | Kowalyszyn, Rubén | Comité Independiente de Etica en investigación clínica "Dr. Carlos A. Barclay, Larrea 1381, 3° A, 1117, Buenos Aires, Ciudad Autónoma de BuenosAires, Argentina | 3/9/15 |
| 279796 | Argentina | Kahl, Susana | Comité de ética en investigación Fundación Oncosalud, Siria 16, 2700, Perga, Buenos Aires, Argentina | 4/16/15 |
| 279795 | Argentina | Varela, Mirta | Comité de Ética Centro de Oncología e Investigación Buenos Aires, Calle 12 #4756, B1880BBF, Berazategui, Buenos Aires, Argentina | 2/1/16 |
| 279794 | Argentina | Kaen, Diego | Comité Independiente de Etica en investigación clínica "Dr. Carlos A. Barclay, Larrea 1381, 3° A, 1117, Buenos Aires, Ciudad Autónoma de BuenosAires, Argentina | 2/19/15 |
| 279793 | Argentina | Picon, Pablo | Comité de Ética Independiente Patagónico, San Martin 391, L6300DVM, Santa Rosa, La Pampa, Argentina | 4/9/15 |
| 278899 | Australia | Singhal, Nimit | Hunter New England Research Ethics and Governance Unit, Locked Bag 1, 2305, New Lambton, New South Wales, Australia | 3/26/15 |
| 278900 | Australia | Gauden, Stan | Tasmania Health and Medical Human Research Ethics Committee, 301 Sandy Bay Road, 7001, Hobart, Tasmania, Australia | 3/30/15 |

| | | | | |
|---|---|---|---|---|
| 278905 | Australia | Crombie, Catherine | Hunter New England Research Ethics and Governance Unit, Locked Bag 1, 2305, New Lambton, New South Wales, Australia | 3/26/15 |
| 278907 | Australia | Blinman, Prunella | Hunter New England Research Ethics and Governance Unit, Locked Bag 1, 2305, New Lambton, New South Wales, Australia | 3/26/15 |
| 278908 | Australia | Potasz, Nicole | Hunter New England Research Ethics and Governance Unit, Locked Bag 1, 2305, New Lambton, New South Wales, Australia | 3/26/15 |
| 280017 | Australia | Lewis, Craig | Hunter New England Research Ethics and Governance Unit, Locked Bag 1, 2305, New Lambton, New South Wales, Australia | 3/26/15 |
| 280019 | Australia | Gill, Sanjeev | Hunter New England Research Ethics and Governance Unit, Locked Bag 1, 2305, New Lambton, New South Wales, Australia | 3/26/15 |
| 281140 | Australia | Eliadis, Paul | Bellberry Human Research Ethics Committee, 123 Glen Osmond Road, 5063, Eastwood, South Australia, Australia | 4/24/15 |
| 280018 | Australia | Richardson, Gary | Cabrini Human Research Ethics Committee, 183 Wattletree Road, 3144, Malvern, Vic, Australia | 7/29/15 |
| 280016 | Australia | Kosmider, Suzanne | Hunter New England Research Ethics and Governance Unit, Locked Bag 1, 2305, New Lambton, New South Wales, Australia | 3/26/15 |
| 278906 | Australia | Millward, Michael | Sir Charles Gairdner Hospital HREC, Hospital Avenue, 6009, Nedlands, Western Australia, Australia | 7/28/15 |
| 278904 | Australia | John, Thomas | Hunter New England Research Ethics and Governance Unit, Locked Bag 1, 2305, New Lambton, New South Wales, Australia | 12/15/15 |
| 278903 | Australia | Joshi, Abhishek | Hunter New England Research Ethics and Governance Unit, Locked Bag 1, 2305, New Lambton, New South Wales, Australia | 3/26/15 |
| 278902 | Australia | Nordman, Ina | Hunter New England Research Ethics and Governance Unit, Locked Bag 1, 2305, New Lambton, New South Wales, Australia | 7/7/15 |
| 278901 | Australia | Boyer, Michael | Sydney Local Health District Human Research Ethics Committee - CRGH, Hospital Road, 2139, Concord, New South Wales, Australia | 6/18/15 |
| 278898 | Australia | O'Byrne, Kenneth | Hunter New England Research Ethics and Governance Unit, Locked Bag 1, 2305, New Lambton, New South Wales, Australia | 3/26/15 |
| 278897 | Australia | Hughes, Brett | Hunter New England Research Ethics and Governance Unit, Locked Bag 1, 2305, New Lambton, New South Wales, Australia | 3/26/15 |

| | | | | |
|---|---|---|---|---|
| 280014 | Austria | Greil, Richard | Ethikkommission für das Bundesland Salzburg, Sebastian-Stief-Gasse 2, 5010, Salzburg, Salzburg, Austria | 10/21/15 |
| 279880 | Austria | Eckmayr, Josef | Ethikkommission des Landes Oberösterreich, Wagner-Jauregg Weg 15, 4020, Linz, , Austria | 10/21/15 |
| 280642 | Belgium | Dirix, Luc | AZ Sint Augustinus, Oosterveldlaan 24, 2610, Wilrijk, Antwerpen, Belgium | 6/16/15 |
| 279755 | Belgium | Collard, Philippe | Comité d’Ethique hospitalo-facultaire Cliniques universitaires Saint-Luc, Promenade de l’Alma 51 bte B1.43.03, 1200, Bruxelles, Brussels, Belgium | 6/16/15 |
| 280979 | Belgium | Jerusalem, Guy | Comité d’Ethique hospitalo-facultaire Cliniques universitaires Saint-Luc, Promenade de l’Alma 51 bte B1.43.03, 1200, Bruxelles, Brussels, Belgium | 6/16/15 |
| 280980 | Belgium | Goeminne, Jean Charles | Comité d’Ethique hospitalo-facultaire Cliniques universitaires Saint-Luc, Promenade de l’Alma 51 bte B1.43.03, 1200, Bruxelles, Brussels, Belgium | 6/16/15 |
| 279754 | Belgium | Alexander, Patrick | Ethisch Comité Werken Glorieux VZW, Glorieuxlaan 55, 9600, Ronse, Oost-Vlaanderen, Belgium | 6/16/15 |
| 283381 | Brazil | Schwartsmann, Gilberto | Comitê de Ética em Pesquisa do Hospital de Clínicas de Porto Alegre, Rua Ramiro Barcelos 2350, 90035-903, Porto Alegre, Rio Grande do Sul, , Brazil | 4/6/16 |
| 283380 | Brazil | Costamilan, Rita de Cassia | Comitê de Ética em Pesquisa da Universidade de Caxias do Sul, Rua Francisco Getúlio Vargas 1130, 95070-560, Caxias do Sul, Rio Grande do Sul, Brazil | 3/31/16 |
| 281359 | Brazil | Brust, Leandro | Comite de Etica em Pesquisa em Seres Humanos do Centro Universitario UNIVATES/RS, Rua Avelino Tallini 171, Bairro Universitário, 95900-000, Lajeado, Rio Grande do Sul, Brazil | 3/1/16 |
| 281357 | Brazil | Abbade Dettino, Aldo | Comitê de Ética em Pesquisa da Fundação Antônio Prudente - AC Camargo Câncer Center, Rua Professor Antonio Prudente 211, 01509-900, São Paulo, São Paulo, Brazil | 6/6/16 |
| 281356 | Brazil | Bruno, Luiz | Comitê de Ética em Pesquisa - Hospital Mãe de Deus, R. José de Alencar 286, 90880-480, Porto Alegre, Rio Grande do Sul, Brazil | 5/6/16 |
| 280820 | Brazil | Campos, Clodoaldo | Comitê de Ética em Pesquisa em Seres Humanos da Irmandade da Santa Casa de Londrina, Rua Espírito Santo, 523, 86010-510, Londrina, Paraná, Brazil | 5/5/16 |
| 280819 | Brazil | Matias, Danielli | Comitê de Ética em Pesquisa em Seres Humanos da Liga Norte Riograndense Contra o Câncer, Rua Dr. Mário Negócio, 2267, 59040-000, Natal, Rio Grande do Norte, Brazil | 2/2/16 |
| 280816 | Brazil | Faccio, Adilson | Comitê de Ética em Pesquisa em Seres Humanos da Universidade de Ribeirão Preto (UNAERP), Avenida Costabile Romano, 2201, Ribeirania, 14096380, Ribeirão Preto, São Paulo, Brazil | 4/13/16 |

| | | | | |
|---|---|---|---|---|
| 280818 | Brazil | Murad, Andre | Comitê de Ética em Pesquisa em Seres Humanos do Hospital Lifecenter, Avenida Do Contorno, 4747, 30110-921, Belo Horizonte, Minas Gerais, Brazil | 2/19/16 |
| 280817 | Brazil | da Silva, Carlos | Comitê de Ética em Pesquisa Fundação Pio XII Hospital de Câncer de Barretos, Rua Antenor Duarte Villela 1331, 14784-400, Barretos, São Paulo, Brazil | 2/18/16 |
| 281353 | Brazil | Tadokoro, Hakaru | Comite de Etica em Pesquisa da Universidade Federal de Sao Paulo - Hospital Sao Paulo, Rua Botucatu 572, 04023-062, São Paulo, , Brazil | 3/17/16 |
| 280815 | Brazil | Aragao, Bruno | Comitê de Ética em Pesquisa em Seres Humanos do Hospital Socor, Rua Tupis, 1540, 30190-062, Belo Horizonte, Minas Gerais, Brazil | 3/9/16 |
| 280142 | Brazil | Girotto, Gustavo | Comitê de Ética em Pesquisa em Seres Humanos da Faculdade de Medicina de São José do Rio Preto, Avenida Brigadeiro Faria Lima 5416, Vila São Pedro, 15090-000, São José Do Rio Preto, , Brazil | 5/26/15 |
| 281203 | Bulgaria | Ilieva, Rumyana | Ethics Committee for Clinical Trials, 5 Sveta Nedelya Square, 1000, Sofia, Sofia-Grad, Bulgaria | 6/17/15 |
| 281201 | Bulgaria | Mihaylova, Zhasmina | Ethics Committee for Clinical Trials, 5 Sveta Nedelya Square, 1000, Sofia, Sofia-Grad, Bulgaria | 1/27/16 |
| 281200 | Bulgaria | Koynov, Krassimir | Ethics Committee for Clinical Trials, 5 Sveta Nedelya Square, 1000, Sofia, Sofia-Grad, Bulgaria | 6/17/15 |
| 281199 | Bulgaria | Dimitrov, Borislav | Ethics Committee for Clinical Trials, 5 Sveta Nedelya Square, 1000, Sofia, Sofia-Grad, Bulgaria | 6/17/15 |
| 288023 | Canada | Comeau, Reginald | Comite d'ethique de la recherche de l' Hopital Maisonneuve-Rosemont, 5415 Boulevard De L'assomption, H1T 2M4, Montreal, Quebec, Canada | 3/17/16 |
| 281190 | Canada | Conter, Henry | William Osler Health system Research Ethics Board, 2100 Bovaird Drive East, L6W 3J7, Brampton, Ontario, Canada | 5/14/15 |
| 281188 | Canada | Whittom, Renaud | Comite d'ethique de la recherche de l' Hopital Maisonneuve-Rosemont, 5415 Boulevard De L'assomption, H1T 2M4, Montreal, Quebec, Canada | 4/7/16 |
| 281186 | Canada | El-Maraghi, Robert | Royal Victoria Regional Health Centre Research Ethics Board, 201 Georgian Drive, L4M 6M2, Barrie, Ontario, Canada | 7/7/15 |
| 281185 | Canada | Dionne, Jean-Luc | Comite d'ethique de la recherche de l' Hopital Maisonneuve-Rosemont, 5415 Boulevard De L'assomption, H1T 2M4, Montreal, Quebec, Canada | 12/21/15 |
| 280021 | Canada | Rothenstein, Jeffrey | Lakeridge Health REB, 1 Hospital Court, L1G 2B9, Oshawa, Ontario, Canada | 7/15/15 |

| | | | | |
|---|---|---|---|---|
| 280053 | Canada | Cournoyer, Ghislain | Centre de Sante et de Services Sociaux de Saint-Jerome (CSSS) - Comite d'ethique de recherche, 290 Rue de Montigny, J7Z 5T3, Saint Jerome, Quebec, Canada | 6/7/16 |
| 291880 | Chile | Yañez Ruiz, Eduardo | Comité Ético-Científico del Servicio de Salud Metropolitano Oriente (CEC-SSMO), Av. Salvador 364, , Santiago, Región-MetropolitanadeSantiago, Chile | 3/29/16 |
| 288021 | Chile | Orellana Ulunque, Eric | Comité de Ética Científica de Clínica Santa María, Avenida Bellavista 0373, 7520379, Santiago, Región-MetropolitanadeSantiago, Chile | 11/9/15 |
| 279942 | Chile | Orlandi Jorquera, Francisco | Comité Ético Científico del Servicio de Salud Metropolitano Oriente (CEC SSMO), Av. Salvador 364, , Santiago, Región-MetropolitanadeSantiago, Chile | 5/26/15 |
| 279941 | Chile | Ahumada Olea, Mónica | Comité Ético Científico y de Investigación Hospital Clinico Universidad de Chile, Santos Dummont 999, 8380456, Santiago, , Chile | 8/5/15 |
| 281316 | France | Gazaille, Virgile | CPP Sud-Est II, Groupement Hospitalier Est- 59 Boulevard Pinel, 69500, Bron, , France | 6/10/15 |
| 281237 | France | Perol, Maurice | CPP Sud-Est II, Groupement Hospitalier Est- 59 Boulevard Pinel, 69500, Bron, , France | 6/10/15 |
| 281235 | France | Huchot, Eric | CPP Sud-Est II, Groupement Hospitalier Est- 59 Boulevard Pinel, 69500, Bron, , France | 6/10/15 |
| 281233 | France | Dixmier, Adrien | CPP Sud-Est II, Groupement Hospitalier Est- 59 Boulevard Pinel, 69500, Bron, , France | 6/10/15 |
| 281232 | France | Corre, Romain | CPP Sud-Est II, Groupement Hospitalier Est- 59 Boulevard Pinel, 69500, Bron, , France | 6/10/15 |
| 281231 | France | Denis, Fabrice | CPP Sud-Est II, Groupement Hospitalier Est- 59 Boulevard Pinel, 69500, Bron, , France | 6/10/15 |
| 281229 | France | Berard, Henri | CPP Sud-Est II, Groupement Hospitalier Est- 59 Boulevard Pinel, 69500, Bron, , France | 6/10/15 |
| 281227 | France | Becht-Rolly, Catherine | CPP Sud-Est II, Groupement Hospitalier Est- 59 Boulevard Pinel, 69500, Bron, , France | 6/10/15 |
| 279929 | France | Moro-Sibilot, Denis | CPP Sud-Est II, Groupement Hospitalier Est- 59 Boulevard Pinel, 69500, Bron, , France | 6/10/15 |
| 279928 | France | Souquet, Pierre Jean | CPP Sud-Est II, Groupement Hospitalier Est- 59 Boulevard Pinel, 69500, Bron, , France | 6/10/15 |

| | | | | |
|---|---|---|---|---|
| 281230 | France | Coudert, Bruno | CPP Sud-Est II, Groupement Hospitalier Est- 59 Boulevard Pinel, 69500, Bron, , France | 6/10/15 |
| 281236 | France | Dayen, Charles | CPP Sud-Est II, Groupement Hospitalier Est- 59 Boulevard Pinel, 69500, Bron, , France | 6/10/15 |
| 291416 | Germany | Frost, Nikolaj | Landesamt für Gesundheit und Soziales Berlin (LaGeSo) Ethik, Fehrbelliner Platz 1, 10707, Berlin, Berlin, Germany | 5/12/16 |
| 281317 | Germany | Dickgreber, Nicolas | Ethik-Kommission der Ärztekammer Westfalen-Lippe und der Medizinischen Fakultät der WWU Munster, Gartenstrasse 210 - 214, 48147, Münster, Nordrhein-Westfalen, Germany | 8/12/15 |
| 280822 | Germany | Reck, Martin | Ethik-Kommissionen bei der Ärztekammer Schleswig-Holstein, Bismarckallee 8-12, 23795, Bad Segeberg, Schleswig-Holstein, Germany | 8/12/15 |
| 280503 | Germany | Engel-Riedel, Walburga | Ethikkommission der Ärztekammer Nordrhein, Tersteegenstrasse 9, 40474, Düsseldorf, Nordrhein-Westfalen, Germany | 8/12/15 |
| 280502 | Germany | Weißinger, Florian | Ethik-Kommission der Ärztekammer Westfalen-Lippe und der Medizinischen Fakultät der WWU Munster, Gartenstrasse 210 - 214, 48147, Münster, Nordrhein-Westfalen, Germany | 8/12/15 |
| 280501 | Germany | Stauder, Heribert | Ethikkommission der Bayerischen Landesärztekammer, Mühlbaurstrasse 16, 81677, München, Bayern, Germany | 8/12/15 |
| 280500 | Germany | Schütte, Wolfgang | Ethik-Kommission der Ärztekammer Sachsen-Anhalt, Am Kirchtor 9, 6108, Halle an der Saale, , Germany | 8/12/15 |
| 279973 | Germany | Wehler, Thomas | Ethik-Kommission bei der Ärztekammer des Saarlandes, Faktoreistrasse 4, 66111, Saarbrücken, Saarland, Germany | 8/12/15 |
| 279972 | Germany | Loges, Sonja | Ethik-Kommission der Ärztekammer Hamburg, Weidestr. 122 b, 22083, Hamburg, Hamburg, Germany | 8/12/15 |
| 279970 | Germany | Lehmann, Markus | Ethik-Kommission der Ärztekammer Westfalen-Lippe und der Medizinischen Fakultät der WWU Munster, Gartenstrasse 210 - 214, 48147, Münster, Nordrhein-Westfalen, Germany | 8/12/15 |
| 279969 | Germany | Kokowski, Konrad | Ethik-Kommission der Bayerischen Landesärztekammer, Mühlbaurstr.16, 81677, München, , Germany | 8/12/15 |
| 279960 | Germany | Schulz, Christian | Ethikkommission an der Universität Regensburg, Landshuter Straße 4, 93047, Regensburg, , Germany | 8/12/15 |
| 279956 | Germany | Sadjadian, Parvis | Ethik-Kommission der Ärztekammer Westfalen-Lippe und der Medizinischen Fakultät der WWU Munster, Gartenstrasse 210 - 214, 48147, Münster, Nordrhein-Westfalen, Germany | 8/12/15 |

| | | | | |
|---|---|---|---|---|
| 279954 | Germany | Rittmeyer, Achim | Ethikkommission der Landesärztekammer Hessen, Im Vogelsgesang 3, 60488, Frankfurt am Main, Hessen, Germany | 8/12/15 |
| 279951 | Germany | Kimmich, Martin | Ethikkommission der Landesärztekammer Baden-Württemberg, Jahnstrasse 40, 70597, Stuttgart, Baden-Württemberg, Germany | 8/12/15 |
| 279950 | Germany | Heinrich, Bernhard | Ethikkommission der Bayerischen Landesärztekammer, Mühlbaurstrasse 16, 81677, München, Bayern, Germany | 8/12/15 |
| 279948 | Germany | Rückert, Anja | Ethikkommission der Landesärztekammer Baden-Württemberg, Jahnstrasse 40, 70597, Stuttgart, Baden-Württemberg, Germany | 8/12/15 |
| 279947 | Germany | Fischer, Jürgen | Ethikkommission der Landesärztekammer Baden-Württemberg, Jahnstrasse 40, 70597, Stuttgart, Baden-Württemberg, Germany | 8/12/15 |
| 279946 | Germany | Behringer, Dirk | Ethik-Kommission der Ärztekammer Westfalen-Lippe und der Medizinischen Fakultät der WWU Munster, Gartenstrasse 210 - 214, 48147, Münster, Nordrhein-Westfalen, Germany | 8/12/15 |
| 279945 | Germany | Bargon, Joachim | Ethikkommission der Landesärztekammer Hessen, Im Vogelsgesang 3, 60488, Frankfurt am Main, Hessen, Germany | 8/12/15 |
| 279944 | Germany | Wesseler, Claas | Ethik-Kommission der Ärztekammer Hamburg, Weidestr. 122 b, 22083, Hamburg, Hamburg, Germany | 8/12/15 |
| 279974 | Germany | Wermke, Martin | Ethik-Kommission der Medizinischen Fakultät "Carl Gustav Carus" der Technischen Universität Dresden, Fetscherstraße 74, 01307, Dresden, , Germany | 8/12/15 |
| 282806 | Germany | Wiewrodt, Rainer | Ethik-Kommission der Ärztekammer Westfalen-Lippe und der Medizinischen Fakultät der WWU Münster, Gartenstrasse 210 - 214, 48147, Münster, Nordrhein-Westfalen, Germany | 8/31/15 |
| 297763 | Germany | Laack, Eckart | Ethik-Kommission der Ärztekammer Westfalen-Lippe und der Medizinischen Fakultät der WWU Münster, Gartenstrasse 210 - 214, 48147, Münster, Nordrhein-Westfalen, Germany | 2/16/17 |
| 281141 | Israel | Kolin, Maya | Galilee Medical Center EC, Western Galilee Hospital POB 21, 22100, Nahariya, , Israel | 5/21/15 |
| 279781 | Israel | Wollner, Mirjana | Rambam Medical Center Ethics Committee, 8 Haaliya Hashniya Street, 31096, Haifa, Haifa, Israel | 7/26/15 |
| 279779 | Israel | Merimsky, Ofer | Tel Aviv Sourasky EC, 6 Weitzman Street, 6423906, TEL AVIV, , Israel | 5/28/15 |
| 279778 | Israel | Bar, Yair | The Chaim Sheba Medical Center EC, Tel Hashomer, 52621, Ramat Gan, , Israel | 4/28/15 |

| | | | | |
|---|---|---|---|---|
| 279777 | Israel | Cyjon, Arnold | Shamir Medical Center Assaf Harofeh EC, Beer Yaakov 70300, 70300, Zerifin, , Israel | 4/16/15 |
| 279775 | Israel | Dudnik, Julia | Soroka University Medical Center Local EC, Reger Avenu, 84101, Beer Sheva, , Israel | 3/5/15 |
| 279774 | Israel | Stemmer, Salomon | Rabin Medical Center Ethics Committee, 39 Jabotinski St., 49100, Petach Tikva, , Israel | 3/29/15 |
| 279773 | Israel | Nechushtan, Hovav | Hadassah University Hospital Local EC, Kiryat Hadassah, 91120, Jerusalem, , Israel | 4/1/15 |
| 279772 | Israel | Katsnelson, Rivka | Kaplan Medical Center Local EC, Kaplan Medical Center, 76100, Rehovot, , Israel | 5/19/15 |
| 279770 | Israel | Gottfried, Maya | Meir EC, 59 Tchernichovsky Street, 44281, Kfar Saba, HaMerkaz, Israel | 6/11/15 |
| 279863 | Italy | Allegrini, Giacomo | Comitato Etico Regionale Toscana – Area Vasta Nord Ovest, Via Roma 67, 56126, Pisa, Toscana, Italy | 7/23/15 |
| 279869 | Italy | Falcone, Alfredo | Comitato Etico Regionale Toscana – Area Vasta Nord Ovest, Via Roma 67, 56126, Pisa, Toscana, Italy | 7/23/15 |
| 279871 | Italy | Gridelli, Cesare | Comitato Etico Campania Nord, VIA DEGLI IMBIMBO 10-12, 83100, Avellino, Camparia, Italy | 6/23/15 |
| 279873 | Italy | Mencoboni, Manlio | Comitato Etico San Martino IST 2, Largo Rosanna Benzi 10, 16132, Genova, , Italy | 9/15/15 |
| 293924 | Italy | Pedrazzoli, Paolo | Comitato di Bioetica Fondazione IRCCS Policlinico S. Matteo di Pavia, Viale Golgi 19, 27100, Pavia, , Italy | 10/10/16 |
| 279875 | Italy | Morabito, Alessandro | Comitato Etico IRCCS<br>Istituto Nazionale per lo Studio e la Cura dei Tumori Fondazione G Pascale, Via Mariano Semmola, 80131, Napoli, Campania, Italy | 8/3/15 |
| 280015 | Italy | Soto-Parra, Hector | Comitato Etico Catania 1, VIA SANTA SOFIA 78, 95123, Catania, Sicilia, Italy | 6/9/15 |
| 281142 | Italy | Minotti, Vincenzo | Comitato Etico delle Aziende Sanitarie dell’Umbria, Via della Rivoluzione, 16, 06070, Ellera di Corciano, Perugia, Italy | 6/30/15 |
| 284676 | Italy | Galetta, Domenico | Comitato Etico Area 5 presso IRCCS Ospedale Oncologico di Bari Istituto Tumori Giovanni Paolo II, Viale Orazio Flacco 65, 70124, Bari, Puglia, Italy | 3/17/16 |

| | | | | |
|---|---|---|---|---|
| 280649 | Italy | Ciardiello, Fortunato | Comitato Etico dell'Università Federico II, Via Sergio Pansini, 5, 80131, Napoli, Napoli, Italy | 7/23/15 |
| 280480 | Italy | Borra, Roberta | Comitato Etico Azienda Ospedaliera Universitaria Maggiore della Carità, Corso Mazzini 18, 28100, Novara, Piemonte, Italy | 8/27/15 |
| 279874 | Italy | Migliorino, Maria | Comitato Etico Lazio 1, Circonvallazione Gianicolense, 87, 152, Roma, Lazio, Italy | 7/30/15 |
| 279866 | Italy | Cartenì, Giacomo | Comitato Etico Cardarelli-Santobono, Via Antonio Cardarelli 9, 80131, Napoli, Campania, Italy | 10/7/15 |
| 279785 | Latvia | Zvirbule, Zanete | The Ethics Committee for Clinical Trials of Medicinal Products, Aizkraukles Street 21-113, LV-1006, Riga, , Latvia | 4/24/15 |
| 279784 | Latvia | Purkalne, Gunta | The Ethics Committee for Clinical Trials of Medicinal Products, Aizkraukles Street 21-113, LV-1006, Riga, , Latvia | 4/24/15 |
| 281218 | Lithuania | Cicenas, Saulius | Lithuanian Bioethics Committee, Vilniaus str. 16, LT-01402, Vilnius, , Lithuania | 5/20/15 |
| 282504 | Mexico | Dominguez Andrade, Adriana | Comite de Etica en Investigacion de Mexico Centre for Clinical Research SA de CV, Amores 709, 3100, Ciudad de México, Distrito Federal, Mexico | 5/15/15 |
| 280831 | Mexico | Noriega Iriondo, Maria | Comité de Ética en Investigación de la Facultad de Medicina de la UANL y Hospital Universitario “Dr., Av. Francisco I. Madero y Gonzalitos S/N, Colonia Mitras Centro, 64460, Monterrey, Nuevo León, Mexico | 8/24/15 |
| 284134 | Netherlands | Dingemans, Anne-Marie | METC azM/UM, Oxfordlaan 10, 6202 AZ, Maastricht, , Netherlands | 10/29/15 |
| 281348 | Netherlands | Snijders, Dominic | METC Noord Holland, Nassauplein 10, 1815 GM, Alkmaar, , Netherlands | 10/29/15 |
| 281347 | Netherlands | Schramel, Franz | Medical Research Ethics Committees United, Koekoekslaan 1, 3435 CM, Nieuwegein, Utrecht, Netherlands | 12/8/15 |
| 279881 | Netherlands | Van den Borne, Ben | Medical Research Ethics Committees United, Koekoekslaan 1, 3435 CM, Nieuwegein, Utrecht, Netherlands | 10/29/15 |
| 279882 | Netherlands | Wilschut, Frank | Medical Research Ethics Committees United, Koekoekslaan 1, 3435 CM, Nieuwegein, Utrecht, Netherlands | 10/29/15 |
| 280825 | Netherlands | Hashemi, Sayed | Medical Research Ethics Committees United, Koekoekslaan 1, 3435 CM, Nieuwegein, Utrecht, Netherlands | 10/29/15 |

| | | | | |
|---|---|---|---|---|
| 281346 | Netherlands | Aerts, Joachim | Medical Research Ethics Committees United, Koekoekslaan 1, 3435 CM, Nieuwegein, Utrecht, Netherlands | 10/29/15 |
| 282503 | Netherlands | Buikhuisen, Wieneke | Medical Research Ethics Committees United, Koekoekslaan 1, 3435 CM, Nieuwegein, Utrecht, Netherlands | 10/29/15 |
| 283733 | Peru | Cisneros Tipismana, Rocio | Comite de Etica en Investigacion del Instituto Regional de Enfermedades Neoplasicas, Panamericana Norte Km. 558, 12345, Trujillo, , Peru | 10/14/15 |
| 283313 | Peru | Moron Escobar, Hernan | Asociacion Benefica Prisma, Calle Carlos Gonzles 251, Lima 32, Lima, Lima, Peru | 5/28/15 |
| 281315 | Peru | Kobashigawa, Alejandro | Comite de Etica en Investigacion del Hospital Guillermo Almenara Irigoyen, Avenida Grau 800, Lima 13, Lima, Lima, Peru | 10/14/15 |
| 279756 | Peru | Aleman Polanco, Diana Sofía | Asociacion Benefica Prisma, Calle Carlos Gonzles 251, Lima 32, Lima, Lima, Peru | 4/23/15 |
| 278307 | Peru | Mas Lopez, Luis | Comité Institucional de ética en Investigación Instituto Nacional de Enfermedades Neoplasicas, Avenida Angamos Este 2520, Lima34, Lima, Lima, Peru | 5/4/15 |
| 279930 | Portugal | Almodovar, Maria Teresa | Comissão de Ética para a Investigação Clínica - CEIC, Avenida do Brasil, 53, 1749-004- Lisboa, Lisboa, Portugal | 7/10/15 |
| 279931 | Portugal | Araújo, Antonio | Comissão de Ética para a Investigação Clínica - CEIC, Avenida do Brasil, 53, 1749-004- Lisboa, Lisboa, Portugal | 7/10/15 |
| 279932 | Portugal | Barata, Fernando | Comissão de Ética para a Investigação Clínica - CEIC, Avenida do Brasil, 53, 1749-004- Lisboa, Lisboa, Portugal | 7/10/15 |
| 279933 | Portugal | Queiroga, Henrique | Comissão de Ética para a Investigação Clínica - CEIC, Avenida do Brasil, 53, 1749-004- Lisboa, Lisboa, Portugal | 7/10/15 |
| 279934 | Portugal | Rodrigues, Ana | Comissão de Ética para a Investigação Clínica - CEIC, Avenida do Brasil, 53, 1749-004- Lisboa, Lisboa, Portugal | 7/10/15 |
| 279935 | Portugal | Teixeira, Encarnação | Comissão de Ética para a Investigação Clínica - CEIC, Avenida do Brasil, 53, 1749-004- Lisboa, Lisboa, Portugal | 7/10/15 |
| 281362 | Russian Federation | Stroyakovskiy, Daniil | Ethics Committee at Moscow City Oncology Hospital #62 of Moscow Healthcare Department, Krasnogorskiy district, Stepanovskoe, settlement Istra, 27, 143423, Moscow, , Russian Federation | 8/14/15 |
| 281361 | Russian Federation | Zhiltsova, Elena | Ethics Committee at Russian Medical Military Academy n.a. S.M.Kirov, Ulitsa Academika Lebedeva, 6, 194044, St. Petersburg, , Russian Federation | 9/22/15 |

| | | | | |
|---|---|---|---|---|
| 281360 | Russian Federation | Gorbunova, Vera | Ethics Committee at Russian Oncology Research Center n.a. N.N.Blokhin, Kashirskoe Shosse 24, 115478, Moscow, , Russian Federation | 9/8/15 |
| 281238 | Russian Federation | Galiulin, Rinat | Ethics Committee at Clinical Oncology Dispensary, Ulitsa Zavertyayeva, 9 - 1, 644013, Omsk, , Russian Federation | 8/4/15 |
| 281239 | Russian Federation | Karaseva, Nina | Ethics Committee at City Clinical oncologic dispensary, Vtoraya Beryozovaya Alleya 3/5, 197022, St. Petersburg, , Russian Federation | 8/4/15 |
| 281363 | Russian Federation | Kovalenko, Nadezhda | Ethics Committee at Volzhskiy regional clinical oncology dispensary #3, Ulitsa Komsomolskaya, 25, 404100, Volzhskiy, , Russian Federation | 6/27/16 |
| 280020 | Singapore | Soo, Ross | Domain Specific Review Board, Nexus@One-North (South Tower), 138543, Singapore, , Singapore | 4/9/15 |
| 278955 | Singapore | Lim, Darren | Singhealth Centralised Institutional Review Board, 7 Hospital Drive, Singhealth Office Of Research, Blk A, #03-01, Singhealth Research Facilities, 169611, Singapore, Singapore, Singapore | 3/10/15 |
| 283382 | Slovakia | Godal, Robert | Eticka komisia pri Narodnom onkologickom ustave, Klenova 1, 833 01, Bratislava, , Slovakia | 6/29/15 |
| 281215 | Slovakia | Kasan, Peter | Eticka komisia Univerzitna nemocnica Bratislava, Ruzinovska 6, 826 06, Bratislava, , Slovakia | 6/29/15 |
| 281213 | Slovakia | Beniak, Juraj | Eticka komisia Presovskeho samospravneho kraja, Namestie Mieru 2, 080 01, Presov, , Slovakia | 6/29/15 |
| 294325 | Spain | Fuentes Pradera, Jose | CEIC de la Corporacion Sanitaria del Parc Tauli, Calle Parc Tauli, s/n, 8208, Sabadell, Barcelona, Spain | 6/28/16 |
| 282505 | Spain | Palmero, Ramón | CEIC Hospital Universitari de Bellvitge, C/ Feixa Llarga s/n, 8907, L´Hospitalet de Llobregat, Cataluña, Spain | 6/9/15 |
| 280982 | Spain | Rodriguez-Abreu, Delvys | CEIC Hospital Universitario Insular Materno-Infantil de Las Palmas, Avenida Marítima del Sur, s/n, 35016, Las Palmas de Gran Canaria, , Spain | 6/9/15 |
| 280981 | Spain | Blasco Cordellat, Ana | CEIC Consorcio Hospital General Universitario de Valencia, Avenida Tres Cruces, 2, 46014, Valencia, Valencia, Spain | 6/9/15 |
| 280829 | Spain | Ponce Aix, Santiago | CEIC Hospital Universitario 12 de Octubre, Avenida de Cordoba, s/n, 28041, Madrid, Madrid, Spain | 6/9/15 |
| 280827 | Spain | Garrido Lopez, Pilar | CEIC Hospital Universitario Ramon y Cajal, Carretera de Colmenar km. 9.100, 28034, Madrid, Madrid, Spain | 6/9/15 |

| | | | | |
|---|---|---|---|---|
| 280826 | Spain | Felip Font, Enriqueta | CEIC Hospital Universitario Vall d'Hebrón, Passeig de la Vall d'Hebron, 119-129, 8035, Barcelona, , Spain | 6/9/15 |
| 280160 | Spain | Viñolas, Nuria | CEIC Hospital Clinic de Barcelona, Calle Villarroel, 170, 8036, Barcelona, Barcelona, Spain | 6/9/15 |
| 280159 | Spain | Vazquez Estevez, Sergio | CEIC de Galicia (CAEI), Edificio Administrativo San Lázaro, s/n, 15781, Santiago de Compostela, A Coruña, Spain | 6/9/15 |
| 280158 | Spain | Terrasa Pons, Josefa | CEIC Islas Baleares (CEIC-IB), Camí de Jesús, 38 A, 7011, Palma de Mallorca, Baleares, Spain | 6/9/15 |
| 280157 | Spain | Taus, Alvaro | CEIC Parc de Salut Mar, Calle Doctor Aiguader, 88, 8003, Barcelona, Barcelona, Spain | 6/9/15 |
| 280156 | Spain | Oramas, Juana | CEIC Hospital Universitario de Canarias, Calle Ofra, s/n - Planta -2, 38320, La Laguna, Santa Cruz de Tenerife, Spain | 6/9/15 |
| 280155 | Spain | Lopez Brea, Marta | CEIC de Cantabria, Avenida Cardenal Herrera Oria, s/n, 39011, Santander, Cantabria, Spain | 6/9/15 |
| 280154 | Spain | Insa Molla, Amelia | CEIC Hospital Clínico Universitario de Valencia, Avenida Vicente Blasco Ibánez, 17, 46010, Valencia, Valencia, Spain | 6/9/15 |
| 280153 | Spain | Jiménez Munarriz, Beatriz | CEIC Grupo Hospital de Madrid, Avenida Monteprincipe, 25, 28660, Boadilla del Monte, Madrid, Spain | 6/9/15 |
| 280152 | Spain | Gonzalez Larriba, Jose Luis | CEIC Hospital Clinico San Carlos, Calle Profesor Martin Lagos, s/n, 28040, Madrid, Madrid, Spain | 6/9/15 |
| 280151 | Spain | Alvarez, Rosa | CEIC Hospital General Universitario Gregorio Marañon, Calle Doctor Esquerdo, 46, 28007, Madrid, Madrid, Spain | 6/9/15 |
| 280150 | Spain | Garcia Campelo, Rosario | CEIC de Galicia (CAEI), Edificio Administrativo San Lázaro, s/n, 15781, Santiago de Compostela, A Coruña, Spain | 6/9/15 |
| 280148 | Spain | Domine Gomez, Manuel | CEIC Fundación Jiménez Díaz, Avenida Reyes Católicos, 2, 28040, Madrid, Madrid, Spain | 6/9/15 |
| 280147 | Spain | De Castro Carpeño, Javier | CEIC Hospital Universitario La Paz, Paseo de la Castellana, 261, 28046, Madrid, Madrid, Spain | 6/9/15 |
| 280146 | Spain | Barneto Aranda, Isidoro Carlos | CEIC de Andalucia (CCEIBA), Avenida de la Innovación s/n, 41020, Sevilla, Andalucía, Spain | 6/9/15 |

| | | | | |
|---|---|---|---|---|
| 280145 | Spain | Artal-Cortes, Angel Fernando | CEIC de Aragon (CEICA), Avenida San Juan Bosco, 13, 50009, Zaragoza, Zaragoza, Spain | 6/9/15 |
| 280144 | Spain | Majem Tarruella, Margarita | CEIC Hospital Santa Creu i Sant Pau, Avenida Sant Antoni Maria Claret, 167, 8025, Barcelona, Barcelona, Spain | 6/9/15 |
| 280143 | Spain | Garcia, Yolanda | CEIC de la Corporacion Sanitaria del Parc Tauli, Calle Parc Tauli, s/n, 8208, Sabadell, Barcelona, Spain | 6/9/15 |
| 280479 | Switzerland | Ochsenbein, Adrian | Kantonale Ethikkommission Bern (KEK), Murtenstraße 31, 3010, Bern, , Switzerland | 3/2/16 |
| 286358 | Taiwan, Province of China | Su, Ying Wen | Mackay Memorial Hospital Institutional Review Board, No.92, Section2, Chung-shan North Road, 104, Taipei, , Taiwan, Province of China | 8/17/15 |
| 280823 | Taiwan, Province of China | Liu, Chien-Ying | Chang Gung Medical Foundation, 199 Tung Hwa North Road, 10507, Taipei, , Taiwan, Province of China | 5/11/15 |
| 280505 | Taiwan, Province of China | Yu, Chong-Jen | Institution Review Board of National Taiwan University Hospital, No.1, Changde-de Street, Zhongzheng Dist, 100, Taipei, , Taiwan, Province of China | 6/22/15 |
| 278964 | Taiwan, Province of China | Chen, Yuh-Min | Institutional Review Board Taipei Veterans General Hospital, No. 201, Sec.2 Shipei Road, Beitou Dist., 11217, Taipei, , Taiwan, Province of China | 6/11/15 |
| 279753 | Taiwan, Province of China | Chen, Wei-Teing | Cheng-Hsin General Hospital Institutional Review Board, 1F, No. 45, Chenghsin Street, Beitou District, 112, Taipei City, , Taiwan, Province of China | 8/11/15 |
| 279752 | Taiwan, Province of China | Ho, Ching-Liang | Institutional Review Board of Tri-Service General Hospital, No.325,Section 2, Cheng-Kung Road, 11490, Taipei, , Taiwan, Province of China | 6/9/15 |
| 279751 | Taiwan, Province of China | Lin, Yu-Ching | Chang Gung Medical Foundation, 199 Tung Hwa North Road, 10507, Taipei, , Taiwan, Province of China | 5/11/15 |
| 279750 | Taiwan, Province of China | Ji, Bin-Chuan | Institutional Review Board, Changhua Christian Hospital, No.135 Nansiao Street, 50006, Changhua, , Taiwan, Province of China | 6/9/15 |
| 278968 | Taiwan, Province of China | Hung, Jen-Yu | Institutional Review Board Kaohsiung Medical University Chung-Ho Memorial Hospital, No.100, Tzyou 1st Road, 807, Kaohsiung City, , Taiwan, Province of China | 6/2/15 |
| 278965 | Taiwan, Province of China | Chang, Gee-Chen | The Institutional Review Board of Taichung Veterans General Hospital, No.160 Section 3 Chung-Kang Road, 40705, Taichung, , Taiwan, Province of China | 6/10/15 |
| 278960 | Taiwan, Province of China | Chen, Chao-Hsun | Institutional Review Board of the Chi Mei Medical Center, 4F, 3rd Medical building, No. 901 Chung-Huwa Rd., Young-Kang Dist. Tainan, Taiwan., , Tainan, , Taiwan, Province of China | 5/20/15 |

| | | | | |
|---|---|---|---|---|
| 278956 | Taiwan, Province of China | Hsia, Te-Chun | China Medical University and Hospital Research Ethics Committee, No.2, Yuh-Der Road, 40447, Taichung, , Taiwan, Province of China | 6/13/15 |
| 280760 | Ukraine | Vynnychenko, Ihor | CEQ of Regional Municipal Institution Sumy Regional Clinical Oncology Dispensary, Vulytsya Pryvokzalna 31, 40005, Sumy, , Ukraine | 3/2/15 |
| 280761 | Ukraine | Kobziev, Oleh | CEQ of Municipal Noncommercial Institution Regional Center of Oncology, Vulytsya Lisoparkivska 4, 61070, Kharkiv, , Ukraine | 3/20/15 |
| 280762 | Ukraine | Vasylyev, Leonid | CEQ of SI Institute of Medical Radiology n.a. S.P. Hryhoriev of NAMS of Ukraine, 82 Pushkinska str., 61024, Kharkiv, , Ukraine | 3/17/15 |
| 280763 | Ukraine | Shparyk, Yaroslav | CEQ of Lviv State Oncology Regional Treatment Diagnostic Center, 2-A Yaroslava Hasheka str., 79031, Lviv, , Ukraine | 3/18/15 |
| 280764 | Ukraine | Rusyn, Andriy | CEQ of Transcarpathian Regional Clinical Oncology Dispensary, Vulytsya Brodlakovycha, 2, 88014, Uzhgorod, , Ukraine | 3/10/15 |
| 280766 | Ukraine | Ivashchuk, Oleksandr | Commission of Ethics Questions on the basis of the Chernivtsi Regional Clinical Oncology Dispensary, Vulytsya Chervonoarmiyska 242, 58013, Chernivtsi, , Ukraine | 5/22/15 |
| 280767 | Ukraine | Hotko, Yevhen | Commission on Ethics Questions of MNPE Central City Clinical Hospital of Uzhhorod City Council, Vulytsya Gryboedova 20, 88000, Uzhgorod, , Ukraine | 3/3/15 |
| 280768 | Ukraine | Chornobai, Anatolii | CEQ of Poltava Regional Clinical Oncology Dispensary of Poltava Regional Council, 7a, Volodarskoho Str., 36021, Poltava, , Ukraine | 2/26/15 |
| 280769 | Ukraine | Bondarenko, Igor | LEC of Municipal Non-profit Enterprise "City Clinical Hospital # 4" of Dnipro City Council, Vulytsya Blyzhnya 31, 49102, Dnipropetrovsk, Dnipropetrovs'ka Oblast , Ukraine | 3/19/15 |
| 280770 | Ukraine | Andrusenko, Orest | CEQ of Treatment and Prevention Institution Volyn Regional Oncology Dispensary, Vulytsya Tymiryazeva 1, 43018, Lutsk, , Ukraine | 5/15/15 |
| 280771 | Ukraine | Adamchuk, Hryhoriy | CEQ of MI Kryvyi Rih Oncology Dispensary of Dnipropetrovsk Regional Council, 41 Dnipropetrovske Road, 50048, Kryvyi Rih, , Ukraine | 2/27/15 |
| 280821 | Ukraine | Goloborodko, Oleksandr | CEQ of MI of Zaporizhzhia Regional Council Zaporizhzhia Regional Clinical Oncology Dispensary, 177-A Kulturna str., 69040, Zaporizhzhia, Zaporiz'ka Oblast, Ukraine | 6/25/15 |
| 288092 | Ukraine | Shamrai, Volodymyr | Commission on Ethics Questions of Vinnytsya Regional Clinical Oncology Dispensary, 84 Khmelnytskyy prospekt, 21029, Vinnytsya, , Ukraine | 8/27/15 |
| 293804 | United States | MacKintosh, Frederick | University of Nevada Reno Biomedical Institutional Review Board, 1664 North Virginia Street, 89557, Reno, Nevada, United States | 7/21/16 |

| | | | | |
|---|---|---|---|---|
| 292662 | United States | Drew, David | Copernicus Group Independent Review Board, 1 Triangle Drive, 27709, Research Triangle Park, North Carolina, United States | 4/12/16 |
| 292661 | United States | Jamil, Rodney | Pinnacle Health Hospitals Institutional Review Board, 205 South Front Street, 17104, Harrisburg, Pennsylvania, United States | 4/12/16 |
| 288792 | United States | Rothschild, Neal | Western Institutional Review Board, 1019 39th Avenue Southeast, 98374, Puyallup, Washington, United States | 10/16/15 |
| 288791 | United States | Suga, Jennifer Marie | Kaiser Permanente Northern California Institutional Review Board, 1800 Harrison Street, 94162, Oakland, California, United States | 6/16/15 |
| 288789 | United States | Suga, Jennifer Marie | Kaiser Permanente Northern California Institutional Review Board, 1800 Harrison Street, 94162, Oakland, California, United States | 6/16/15 |
| 288788 | United States | Suga, Jennifer Marie | Kaiser Permanente Northern California Institutional Review Board, 1800 Harrison Street, 94162, Oakland, California, United States | 6/16/15 |
| 288787 | United States | Suga, Jennifer Marie | Kaiser Permanente Northern California Institutional Review Board, 1800 Harrison Street, 94162, Oakland, California, United States | 6/16/15 |
| 288786 | United States | Suga, Jennifer Marie | Kaiser Permanente Northern California Institutional Review Board, 1800 Harrison Street, 94162, Oakland, California, United States | 6/16/15 |
| 288785 | United States | Suga, Jennifer Marie | Kaiser Permanente Northern California Institutional Review Board, 1800 Harrison Street, 94162, Oakland, California, United States | 6/16/15 |
| 288784 | United States | Suga, Jennifer Marie | Kaiser Permanente Northern California Institutional Review Board, 1800 Harrison Street, 94162, Oakland, California, United States | 6/16/15 |
| 288783 | United States | Suga, Jennifer Marie | Kaiser Permanente Northern California Institutional Review Board, 1800 Harrison Street, 94162, Oakland, California, United States | 6/16/15 |
| 288782 | United States | Sullivan, Kevin | Western Institutional Review Board, 1019 39th Avenue Southeast, 98374, Puyallup, Washington, United States | 3/28/16 |
| 288781 | United States | Fabregas, Jesus | Copernicus Group Independent Review Board, 1 Triangle Drive, 27709, Research Triangle Park, North Carolina, United States | 10/21/15 |
| 288020 | United States | Patel, Pareshkumar | Western Institutional Review Board, 1019 39th Avenue Southeast, 98374, Puyallup, Washington, United States | 9/11/15 |
| 288018 | United States | Ikpeazu, Chukwuemeka | University Of Miami, 1500 N.w. 12th Avenue, 33136, Miami, Florida, United States | 12/1/15 |

| | | | | |
|---|---|---|---|---|
| 287057 | United States | Johns, Mark | US Oncology Inc. Institutional Review Board, 10101 Woodloch Forest, 77380, The Woodlands, Texas, United States | IEC since 30/Sep/2016 |
| 287057 | United States | Johns, Mark | Western Institutional Review Board, 1019 39th Avenue Southeast, 98374, Puyallup, Washington, United States | 9/29/15 |
| 287056 | United States | Daniel, Davey | Western Institutional Review Board, 1019 39th Avenue Southeast, 98374, Puyallup, Washington, United States | 9/3/15 |
| 287054 | United States | Arnaoutakis, Konstantinos | University of Arkansas IRB, 4301 W. Markham Street, 72205, Little Rock, Arkansas, United States | 6/29/16 |
| 287050 | United States | Alnsour, Mohammad | Mercy Saint Vincent Medical Center Institutional Review Board, 2213 Cherry Street, 43608, Toledo, Ohio, United States | 12/15/15 |
| 287048 | United States | Page, Ray | Western Institutional Review Board, 1019 39th Avenue Southeast, 98374, Puyallup, Washington, United States | 9/11/15 |
| 284314 | United States | Spira, Alexander | US Oncology Inc. Institutional Review Board, 10101 Woodloch Forest, 77380, The Woodlands, Texas, United States | 6/18/15 |
| 284312 | United States | Narang, Mohit | US Oncology Inc. Institutional Review Board, 10101 Woodloch Forest, 77380, The Woodlands, Texas, United States | 6/18/15 |
| 284133 | United States | Cetnar, Jeremy | Oregon Health & Science University IRB, 3181 S.W. Sam Jackson Park Road, 97239-3098, Portland, Oregon, United States | 2/12/16 |
| 284132 | United States | Socoteanu, Matei | US Oncology Inc. Institutional Review Board, 10101 Woodloch Forest, 77380, The Woodlands, Texas, United States | 6/18/15 |
| 283311 | United States | Herman, James | WIRB Copernicus Group, 1 Triangle Drive, 27709, Research Triangle Park, North Carolina, United States | 2/18/16 |
| 283308 | United States | Silberberg, Jeffrey | Copernicus Group Independent Review Board, 1 Triangle Drive, 27709, Research Triangle Park, North Carolina, United States | 5/28/15 |
| 283306 | United States | Nissenblatt, Michael | Copernicus Group Independent Review Board, 1 Triangle Drive, 27709, Research Triangle Park, North Carolina, United States | 6/1/15 |
| 283305 | United States | McCleod, Michael | Western Institutional Review Board, 1019 39th Avenue Southeast, 98374, Puyallup, Washington, United States | 9/4/15 |
| 283286 | United States | Hussein, Maen | Western Institutional Review Board, 1019 39th Avenue Southeast, 98374, Puyallup, Washington, United States | 9/11/15 |

| | | | | |
|---|---|---|---|---|
| 283284 | United States | Gurubhagavatula, Sarada | Copernicus Group Independent Review Board, 1 Triangle Drive, 27709, Research Triangle Park, North Carolina, United States | 5/28/15 |
| 282386 | United States | Wender, Donald | Siouxland Institutional Review Board, 230 Nebraska Street, 51101, Sioux City, Iowa, United States | 8/12/15 |
| 282385 | United States | Hoffman, Philip | University of Chicago Hospitals Institutional Review Board, 5751 South Woodlawn Avenue, 60637, Chicago, Illinois, United States | 8/4/15 |
| 282383 | United States | Gunturu, Krishna | Lahey Clinic, Inc. Institutional Review Board, 41 Mall Road, 1805, Boston, Massachusetts, United States | 3/9/16 |
| 282382 | United States | Seng, Sonia | New England Institutional Review Board, 85 Wells Avenue, 2459, Newton, Massachussets, United States | 3/31/16 |
| 282381 | United States | Hooberman, Arthur | Copernicus Group Independent Review Board, 1 Triangle Drive, 27709, Research Triangle Park, North Carolina, United States | 4/20/15 |
| 281314 | United States | Paschold, John | US Oncology Inc. Institutional Review Board, 10101 Woodloch Forest, 77380, The Woodlands, Texas, United States | 6/18/15 |
| 281313 | United States | Knapp, Mark | Copernicus Group Independent Review Board, 1 Triangle Drive, 27709, Research Triangle Park, North Carolina, United States | 6/15/15 |
| 280978 | United States | Sumrall, Bradley | Copernicus Group Independent Review Board, 1 Triangle Drive, 27709, Research Triangle Park, North Carolina, United States | 7/9/15 |
| 280977 | United States | Montero, Aldemar | Copernicus Group Independent Review Board, 1 Triangle Drive, 27709, Research Triangle Park, North Carolina, United States | 3/31/15 |
| 280977 | United States | Montero, Aldemar | Western Institutional Review Board, 1019 39th Avenue Southeast, 98374, Puyallup, Washington, United States | IEC since 24-Sep-2015 |
| 280976 | United States | Matrana, Marc | Ochsner Clinic Foundation Institutional Review Board, 1514 Jefferson Highway, 70121, New Orleans, Louisiana, United States | 5/12/15 |
| 280975 | United States | Koh, Han | Kaiser Permanente Southern California Institutional Review Board., 393 E. Walnut, 91188, Pasadena, California, United States | 5/19/15 |
| 280974 | United States | Kellum, Andrew | North Mississippi Health Services, 830 South Gloster Street, 38801, Tupelo, Mississippi, United States | 5/21/15 |
| 280973 | United States | Goueli, Basem | Saint Luke's Hospital Institutional Review Board, 915 East First Street, 55805, Duluth, Minnesota, United States | 9/1/15 |

| | | | | |
|---|---|---|---|---|
| 280972 | United States | Cho, Jonathan | Copernicus Group Independent Review Board, 1 Triangle Drive, 27709, Research Triangle Park, North Carolina, United States | 3/26/15 |
| 280971 | United States | Nikolinakos, Petros | Copernicus Group Independent Review Board, 1 Triangle Drive, 27709, Research Triangle Park, North Carolina, United States | 3/20/15 |
| 280185 | United States | Jotte, Robert | US Oncology Inc. Institutional Review Board, 10101 Woodloch Forest, 77380, The Woodlands, Texas, United States | 6/18/15 |
| 280181 | United States | Desai, Meghna | Springfield Committee for Research Involving Human Subjects (SCRIHS), 801 North Rutledge Street, 62702, Springfield, Illinois, United States | 11/11/15 |
| 280179 | United States | Rich (Thompson), Patricia | Western Institutional Review Board, 1019 39th Avenue Southeast, 98374, Puyallup, Washington, United States | 10/17/15 |
| 280177 | United States | Thomas, Christian | Copernicus Group Independent Review Board, 1 Triangle Drive, 27709, Research Triangle Park, North Carolina, United States | 5/8/15 |
| 280176 | United States | Stella, Philip | St. Joseph Mercy Health System Institutional Review Board #2 - Oncology Central IRB, 5301 East Huron River Drive, 48106, Ann Arbor, Michigan, United States | 12/18/15 |
| 280174 | United States | McCune, Steven | Copernicus Group Independent Review Board, 1 Triangle Drive, 27709, Research Triangle Park, North Carolina, United States | 10/1/15 |
| 280172 | United States | Nagasaka, Misako | Western Institutional Review Board, 1019 39th Avenue Southeast, 98374, Puyallup, Washington, United States | 7/10/15 |
| 280171 | United States | Finley, Gene | Copernicus Group Independent Review Board, 1 Triangle Drive, 27709, Research Triangle Park, North Carolina, United States | 6/16/15 |
| 280169 | United States | Suga, Jennifer Marie | Kaiser Permanente Northern California Institutional Review Board, 1800 Harrison Street, 94162, Oakland, California, United States | 6/16/15 |
| 280164 | United States | Czerlanis, Cheryl | Loyola University Institutional Review Board, 2160 South First Avenue, 60153, Maywood, Illinois, United States | 6/30/16 |
| 280163 | United States | Goodman, Michael | W.G. 'Bill' Hefner VA Medical Center, 1601 Brenner Avenue, 28144, Salisbury, North Carolina, United States | 3/23/16 |
| 280162 | United States | Kundra, Ajay | Western Institutional Review Board, 1019 39th Avenue Southeast, 98374, Puyallup, Washington, United States | 4/15/15 |
| 280161 | United States | Cohenuram, Michael | Copernicus Group Independent Review Board, 1 Triangle Drive, 27709, Research Triangle Park, North Carolina, United States | 3/13/15 |

| | | | | |
|---|---|---|---|---|
| 279749 | United States | Chitneni, Shobha | Copernicus Group Independent Review Board, 1 Triangle Drive, 27709, Research Triangle Park, North Carolina, United States | 1/29/15 |
| 279748 | United States | Tsai, Frank Yung-Chin | Western Institutional Review Board, 1019 39th Avenue Southeast, 98374, Puyallup, Washington, United States | 7/8/15 |
| 279747 | United States | Swanson, Paul | Copernicus Group Independent Review Board, 1 Triangle Drive, 27709, Research Triangle Park, North Carolina, United States | 2/20/15 |
| 279746 | United States | Subramanian, Janakiraman | Saint Luke's Hospital Institutional Review Board, 4401 Wornall Road, 64111, Kansas City, Missouri, United States | 6/8/15 |
| 279745 | United States | Mitchell, Reed | Copernicus Group Independent Review Board, 1 Triangle Drive, 27709, Research Triangle Park, North Carolina, United States | 1/27/15 |
| 279744 | United States | Kirshner, Eli | Western Institutional Review Board, 1019 39th Avenue Southeast, 98374, Puyallup, Washington, United States | 5/14/15 |
| 279743 | United States | Halibey, Bohdan | Copernicus Group Independent Review Board, 1 Triangle Drive, 27709, Research Triangle Park, North Carolina, United States | 5/29/15 |
| 279742 | United States | Goldschmidt, Jerome | Copernicus Group Independent Review Board, 1 Triangle Drive, 27709, Research Triangle Park, North Carolina, United States | 2/18/15 |
| 279741 | United States | DeVore, Russell | Copernicus Group Independent Review Board, 1 Triangle Drive, 27709, Research Triangle Park, North Carolina, United States | 6/3/15 |
| 279740 | United States | Chaudhry, Arvind | Copernicus Group Independent Review Board, 1 Triangle Drive, 27709, Research Triangle Park, North Carolina, United States | 1/30/15 |
| 279739 | United States | Belman, Neil | St. Luke’s Hospital & Health Network IRB, 801 Ostrum Street, 18015, Bethlehem, Pennsylvania, United States | 3/17/15 |
| 279736 | United States | Almubarak, Mohammed | Advarra Institutional Review Board, 6940 Columbia Gateway Drive, 21046, Columbia, Maryland, United States | 8/13/15 |
| 279735 | United States | Morris, John | Western Institutional Review Board, 1019 39th Avenue Southeast, 98374, Puyallup, Washington, United States | 2/19/16 |
| 279732 | United States | Kerr, Samuel | Lancaster General Hospital IRB, 555 North Duke Street, 17604, Lancaster, Pennsylvania, United States | 3/26/15 |
| 279731 | United States | Bailey, Samuel | Copernicus Group Independent Review Board, 1 Triangle Drive, 27709, Research Triangle Park, North Carolina, United States | 12/11/15 |

| | | | | |
|---|---|---|---|---|
| 279730 | United States | Coleman, Morton | Copernicus Group Independent Review Board, 1 Triangle Drive, 27709, Research Triangle Park, North Carolina, United States | 5/19/15 |
| 279729 | United States | Brzezniak, Christina | Walter Reed National Military Medical Center IRB, 503 Robert Grant Avenue, 20910-7500, Silver Spring, Maryland, United States | 1/7/16 |
| 279727 | United States | Ali, Muhammad | Copernicus Group Independent Review Board, 1 Triangle Drive, 27709, Research Triangle Park, North Carolina, United States | 4/3/15 |
| 278280 | United States | Shtivelband, Mikhail | Copernicus Group Independent Review Board, 1 Triangle Drive, 27709, Research Triangle Park, North Carolina, United States | 6/8/15 |
| 278279 | United States | Erickson, Brian | Copernicus Group Independent Review Board, 1 Triangle Drive, 27709, Research Triangle Park, North Carolina, United States | 2/25/15 |
| 278278 | United States | Burhani, Nafisa | Copernicus Group Independent Review Board, 1 Triangle Drive, 27709, Research Triangle Park, North Carolina, United States | 1/30/15 |
| 278274 | United States | Hamm, John | Western Institutional Review Board, 1019 39th Avenue Southeast, 98374, Puyallup, Washington, United States | 6/2/15 |
| 278272 | United States | Beck, Joseph | Copernicus Group Independent Review Board, 1 Triangle Drive, 27709, Research Triangle Park, North Carolina, United States | 1/7/15 |
| 278271 | United States | Sadiq, Ahad | Copernicus Group Independent Review Board, 1 Triangle Drive, 27709, Research Triangle Park, North Carolina, United States | 3/18/15 |
| 279734 | United States | Wilks, Sharon | Copernicus Group Independent Review Board, 5000 CentreGreen Way, 27513, Cary, North Carolina, United States | 2/9/15 |
| 279768 | United States | Martin, William | St Charles Medical Center, 2500 Northeast Neff Road, 97701, Bend, Oregon, United States | 5/7/15 |
| 288790 | United States | Suga, Jennifer Marie | Kaiser Permanente Northern California Institutional Review Board, 1800 Harrison Street, 94162, Oakland, California, United States | 6/16/15 |
